# Comparison of adverse events between COVID-19 and Flu vaccines

**DOI:** 10.1101/2021.09.22.21263711

**Authors:** Prajwal Mani Pradhan, Zhoutian Shen, Changye Li, Michael J. Remucal

## Abstract

**BACKGROUND:** Among the various driving factors for vaccine hesitancy, confidence in the safety associated with the vaccine constitutes as one of the key factors. This study aimed at comparing the adverse effects of COVID-19 vaccines with the Flu vaccines.

**METHODS:** The VAERS data from 01/01/2020 to 08/20/2021 were used. The MedDRA terms coded by VAERS were further aggregated by a clinician into clinically meaningful broader terms.

**RESULTS:** Various common adverse events between Flu and COVID-19 vaccines have been identified. Adverse events such as headache and fever were very common across all age groups. Among the common adverse events between Flu and COVID-19 vaccine, the relative risk along with 95% CI indicated that such common adverse events were more likely to be experienced by COVID-19 vaccine users than Flu vaccine users. Our study also quantified the proportion of rare adverse events such as Guillain Barre Syndrome and Gynecological changes in the VAERS database for COVID-19 vaccines.

**CONCLUSIONS:** Based on the available data and results, it appears that there were some common adverse events between Flu vaccines and COVID-19 vaccines. These identified common adverse events warrant further investigations based on the relative risk and 95% CI.

## 1. Introduction

The severe acute respiratory syndrome coronavirus 2 (SARS-CoV-2) and its associated pandemic-causing disease, coronavirus disease 2019 (COVID-19) are at the center of the largest public health crisis in over a century. As of the beginning of September 2021, it has resulted in over 4.5 million deaths worldwide and has involved over 215 million people[1].

Considering the serious public health risk posed by the pandemic, the Food and Drug Administration (FDA) issued three Emergency Use Authorizations (EUA) on COVID-19 vaccines. The first Emergency Use Authorization (EUA) issued December 11, 2020, was for the Pfizer-BioNTech Coronavirus disease 2019 (COVID-19) messenger RNA (mRNA) vaccine (BNT162b2) and was indicated for the prevention of COVID-19 among individuals aged 16 years and above[2]. This EUA was further expanded on May 10, 2021, for use in patients 12 years and above[2]. Similarly, the FDA issued a second EUA for the Moderna COVID-19 mRNA vaccine (mRNA-1273) indicated for the prevention of COVID-19 among individuals aged 18 years and older on December 18, 2020 [3]. Finally, the FDA issued a EUA for the Janssen COVID-19 attenuated viral vector vaccine (Ad26CoV2S) indicated for the prevention of COVID-19 among individuals aged 18 years and older on February 27, 2021[4]. Although the Janssen vaccine was paused on April 13^th^, 2021 due to six reports of a rare and severe type of blood clot following vaccination, the pause was lifted on April 23^rd^, 2021 after a thorough safety review by the Centers for Disease Control (CDC) and FDA[5,6].

Despite the emergency approval and quick rollout of COVID-19 vaccines, the uptake of the COVID-19 vaccines has been uneven across various parts of the United States (U.S.)[7]. A study estimated U.S. COVID-19 vaccine hesitancy[8] to be around 22% [9], although a recent CDC report also has suggested that vaccine hesitancy has been decreasing to 16% [10]. Another study estimated that 3 in 10 adults were not sure about their acceptance of vaccination and 1 in 10 did not intend to be vaccinated against COVID-19 [11]. Vaccine hesitancy reduces the impact of COVID-19 vaccination and exacerbates existing disparities in COVID-19 health outcomes [12]. Therefore, to avoid this unfavorable outcome, the government has incentivized the uptake of COVID-19 vaccine [13,14]. However, multiple studies characterize vaccine hesitancy as a multi-layer problem [15,16] which requires a multi-faceted approach to address. Some of the common COVID-19 vaccine hesitancy reasons are political affiliations [15], safety, and concerns of vaccine side effects [12,17,18], trust-related with information from public health experts [17], evidence-based communications; accelerated vaccine or haste of vaccine development [16,19], and concerns around the novelty of available vaccines [20]. A U.S. government report claimed that fear of side effects was the most commonly cited reason for vaccine hesitancy [10]. The same report also cited that the pause on Janssen COVID-19 vaccine impacted some demographic groups more than others [21].

On the other hand, Flu vaccines have been around for more than 75 years and are widely trusted and considered safe. Under such circumstances, it is natural for the public and clinicians to compare the safety profiles, including Adverse Events (AEs), between COVID-19 and Flu vaccines. While several studies are being conducted to monitor the adverse events of COVID-19 vaccines prospectively. Based on our literature review, this is the first study that compares the three popular COVID-19 vaccines (Pfizer, Moderna, and Janssen) and flu vaccine’s AE using an observational study design with the VAERS database.

This study first aimed to compare the top 10 common adverse events of COVID-19 vaccines with the Flu vaccines among persons who reported at least the first dose of receiving COVID-19 or Flu vaccine as reported to VAERS database. Among those top 10 common adverse events, relative risk and associated 95% confidence intervals were calculated as a measure of association. These comparisons will be useful to address the evidence gap in communicating the risk associated with COVID-19 vaccination and thus overcome one of the component of vaccine hesitancy.

## 2. Methods

### 2.1 Data sources

VAERS is a passive national surveillance system administered by the CDC, the FDA, and agencies of the Department of Health and Human Services (DHHS) [22]. While anyone can report adverse effect information to VAERS, health care providers and vaccine manufacturers have certain obligations to report a subset or all adverse effects that they become aware of to VAERS. Adverse event reports in the VAERS database are indicative of a potential association between vaccine and adverse events but these associations should not be implied as causal associations [23]. As with any passive surveillance system it is also susceptible to underreporting and incomplete data [23]. However, the VAERS database can help in identifying safety signals that may indicate potential safety issues which can lead to further detailed examination. Reported adverse events from each report were coded in the VAERS using Medical Dictionary for Regulatory Activities (MedDRA) [23].

### 2.2 Study Design and Cohort

The VAERS data from 01/01/2020 to 08/20/2021 were used for this descriptive study. Additional eligibility criteria were an age of 12 years or older and an indication of either COVID-19 or Flu vaccination in the data. Records with missing age data, or records where a manufacturer of the first vaccine dose could not be ascertained (e.g., cases where information was only available on the second dose of the vaccine) and records that indicated use of COVID-19 vaccines from two different manufacturers (e.g., cases where person received Moderna for their first dose and Pfizer for their second dose) were excluded. A study flow diagram illustrating cohort sizes is shown in Figure 1.

**Figure 1:**
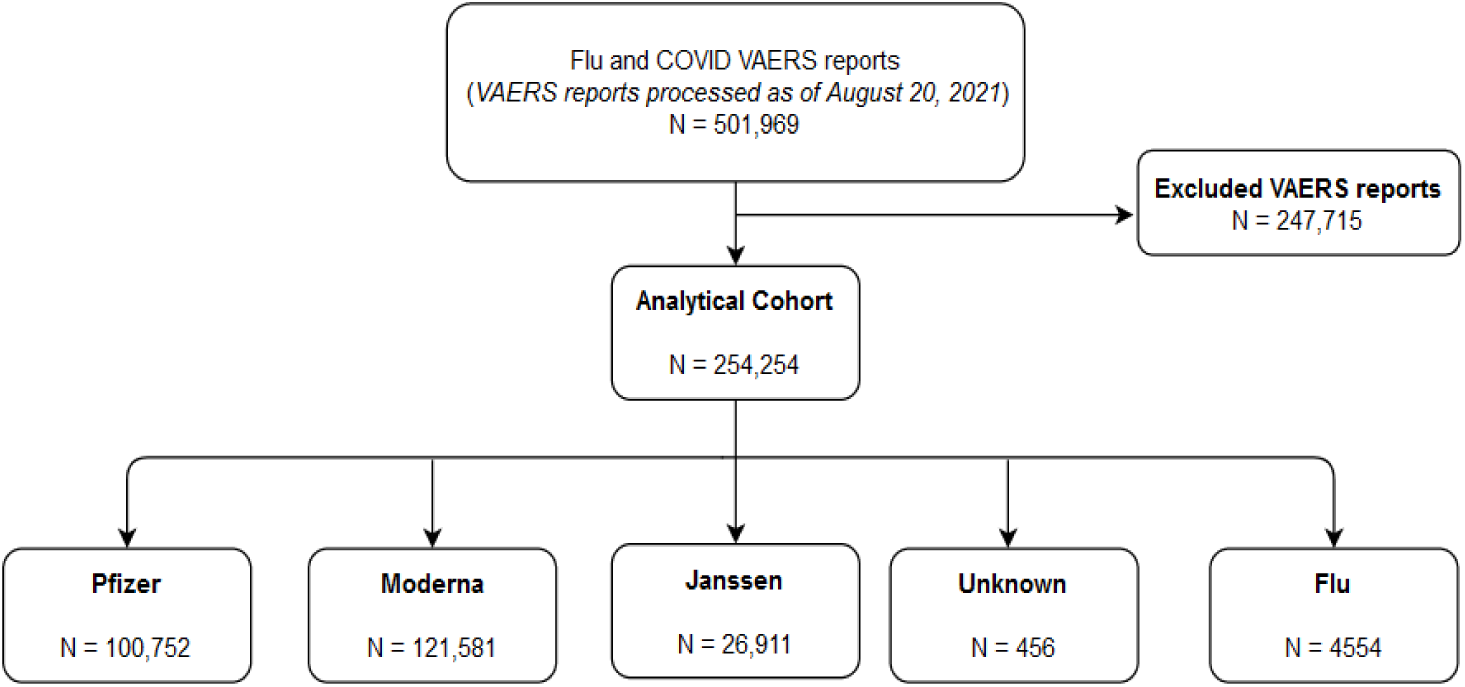
Study Flow diagram representing the cohort.

### 2.3 Data Preprocessing and Outcomes

The MedDRA terms coded by VAERS were further aggregated by a clinician into clinically meaningful broader terms. The reported adverse events in the results section used these broader terms. Mapping of MedDRA terms to broader categories are included in the Supplementary table (**Supplementary table-S1**). All preprocessing of the data was informed by the VAERS Data Use Guide [23].

### 2.4 Statistical Analysis

Demographic information and pregnancy characteristics were described for persons who reported adverse events related to COVID-19 and Flu vaccine use. After stratification of the data based on age-group, we gathered a list of Adverse Events and then reported the top 10 most frequently reported adverse events for each age group and vaccine type. The top 10 AE for each age group and vaccine type are presented in Table 2 whereas, full list of reported adverse events have been included as supplementary tables. Given the intuitive interpretation and other benefits of risk ratios[24,25], relative risks and associated 95 % confidence intervals (CI) were calculated for top 10 AE for each age group and vaccine type.

Data preprocessing and descriptive analyses were conducted using R version 4.0.2, running in RStudio Version 1.2.5033.

## 3. Results

After the application of the methods described above for the study period of 01/01/2020 to 08/20/2021, Moderna had the highest number of adverse event reports (121,581), followed by Pfizer (100,752), Janssen (26,911) and Unknown COVID vaccine (456). For the same study period, there were 4,554 reports associated with the Flu vaccine. A majority of reports were issued on female patients across all five study cohorts: Pfizer (70.6%), Moderna (75.5%), Janssen (63.3%), Unknown COVID vaccine (67.5%) and Flu vaccine (72.0%). In addition, most reports across the five study cohorts were associated with the age group of 31-64 years, Pfizer (59.2%), Moderna (56.1%), Janssen (64.5%), Unknown COVID vaccine (62.5%) and Flu vaccine (41.7%). Across the five cohorts more than 56% of Pfizer, Janssen, Unknown COVID vaccine and Flu vaccine reports had missing information on type of medical attention received (Table 1).

**Table 1:**
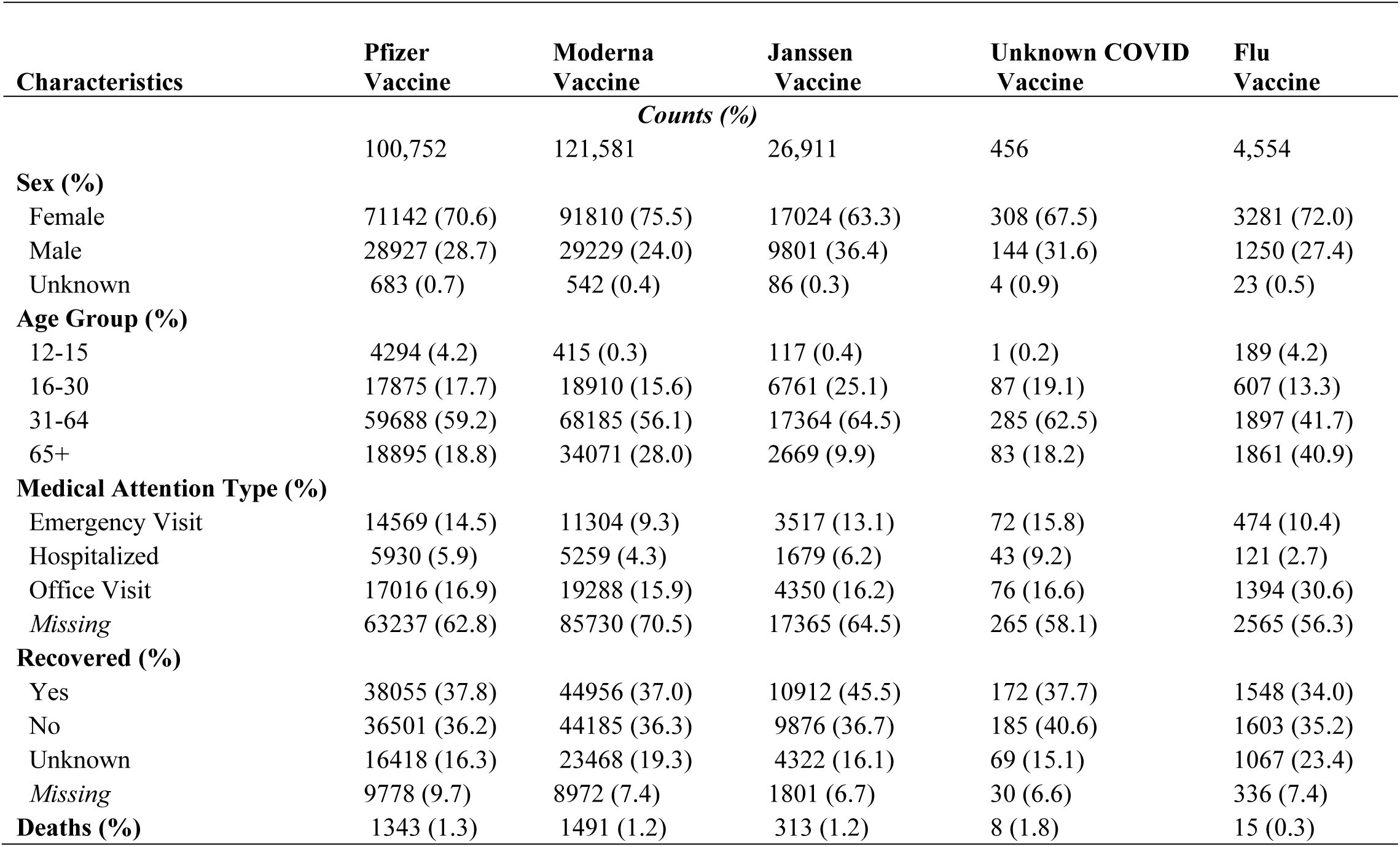
Characteristics of persons who reported use of Flu or COVID Vaccines during 2020 and 2021.

Across the five cohorts, over a third of reports indicated recovery from the associated adverse effects. However, another third of reports indicated that they had not recovered from the vaccine-related adverse effects. The proportion of reported death was at similar level across all COVID-19 vaccines. On the other hand, Flu cohort had a reported death of 0.3% (Table 1). For all five cohorts (i.e. Pfizer, Moderna, Janssen, Unknown COVID-19 and Flu Vaccine cohorts), majority of reports originated in geographical location from the state of California for all five cohorts (***Supplemental table-S2***).

### 3.1 Common adverse events for COVID-19 and Flu Vaccines

The common vaccine manufacturers reported in the data differed by age groups. For the 12 to 15-year age group, Pfizer had the largest number of reports compared to other four cohorts. The most frequently reported adverse events for the Pfizer vaccine in this age group were central neuropathy (24%) for the first dose, and chest pain (31%) for the second dose. The most frequently reported adverse events for the Moderna vaccine were injection site complication (1%), fever (1%), nonspecific musculoskeletal pain (1%) for the first dose and headache (16%), nosebleed (16%), and weakness (16%) for the second dose. The most frequently reported adverse event for the Janssen vaccine was headache (4%) for the first dose. A single adverse event record for the Unknown COVID Vaccine reported having chest pain. In comparison, the most frequently reported adverse event for the Flu vaccine was hypotension (32%) for the first dose. At this age group, the most common adverse events across the five vaccine cohorts were central neuropathy, fever, headache, chest pain, hypotension (Table 2).

**Table 2:**
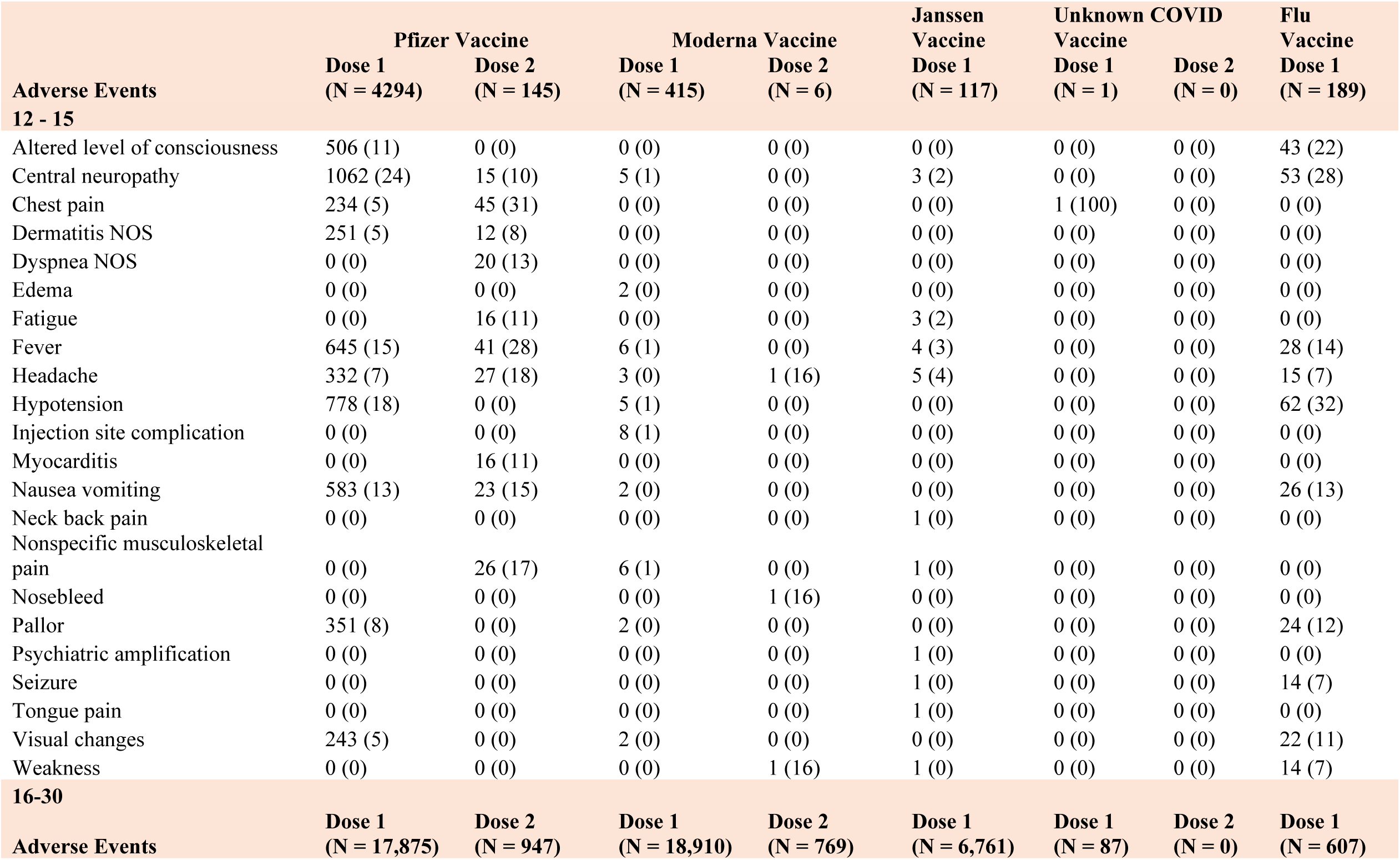
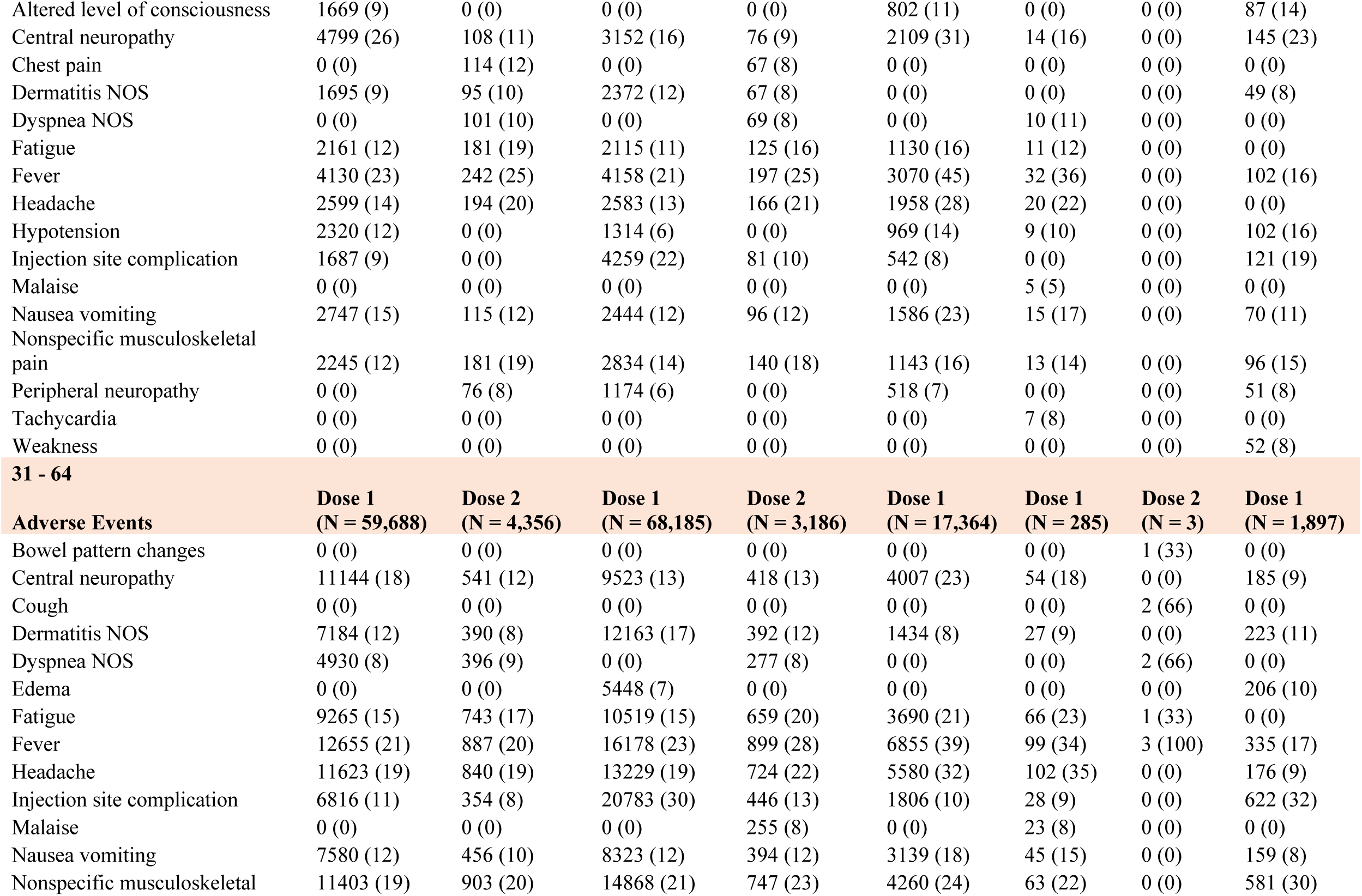
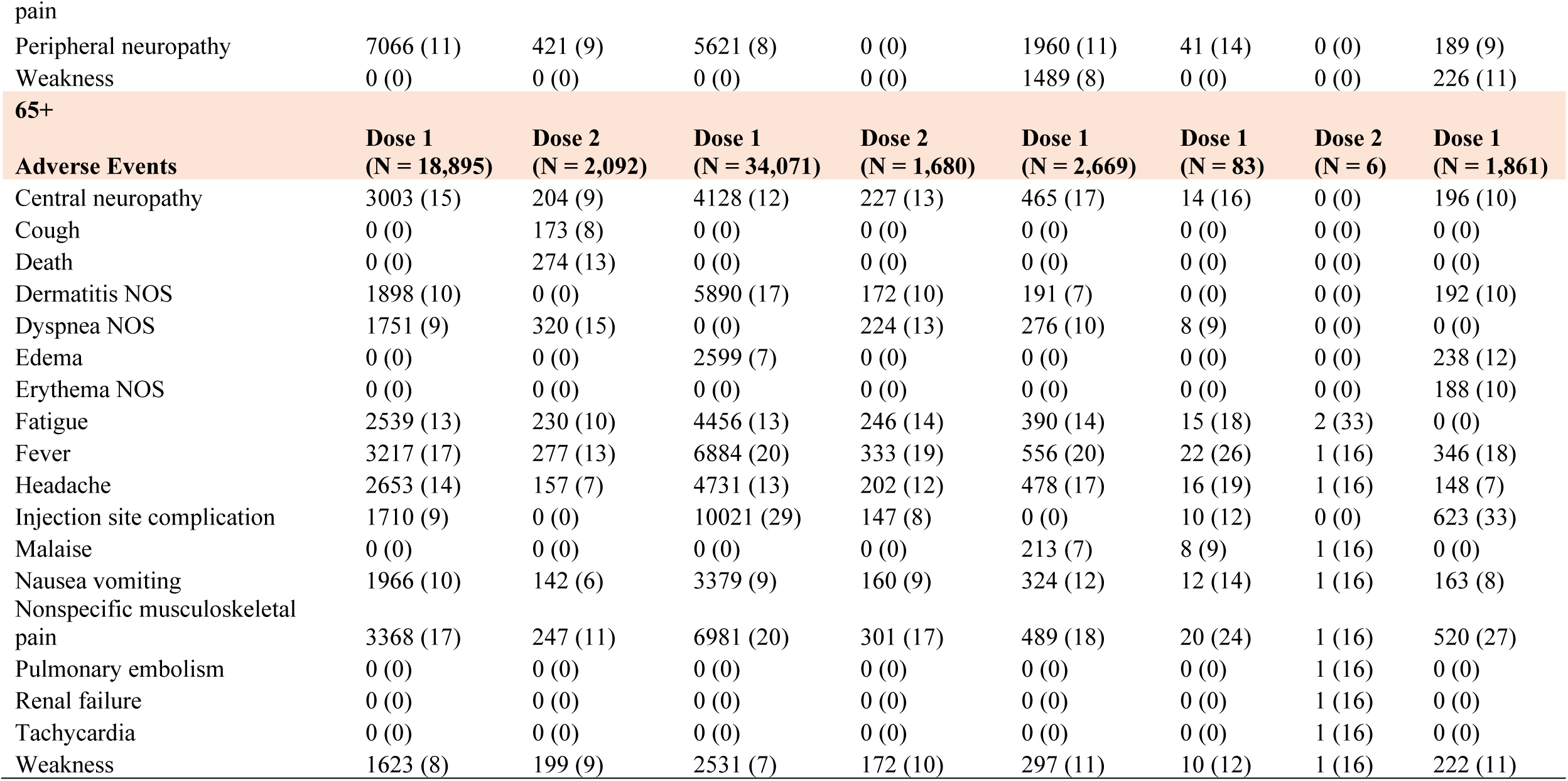
Top 10 frequently reported Adverse events by manufacturer and age groups (*presented in Alphabetical order*)

For the 16 to 30-year age group, Moderna had the largest number of reports compared to other four cohorts. The most frequently reported adverse event for the Pfizer vaccine in this age group were central neuropathy (26%) for the first dose and fever (25%) for the second dose. The most frequently reported adverse events for the Moderna vaccine were injection site complication (22%) for the first dose and fever (25%) for the second dose. The most frequently reported adverse event for Janssen vaccine was fever (45%) for the first dose. The most frequently reported adverse event for the Unknown COVID vaccine was Fever (36%) for the first dose. The most frequently reported adverse event for the Flu vaccine was central neuropathy (23%). At this age group, the most common adverse events across the five-vaccine cohort were fever, central neuropathy, and injection site complication (Table 2).

For the 31 to 64-year age group, Moderna had the largest number of reports compared to other four cohorts. The most frequently reported adverse events for the Pfizer vaccine were fever (21%) for the first dose, and fever (20%) and nonspecific musculoskeletal pain (20%) for the second dose. The most frequently reported adverse events for the Moderna vaccine were injection site complication (30%) for the first dose and fever (28%) for the second dose. The most frequently reported adverse event for the Janssen vaccine was Fever (39%) for the first dose. The most frequently reported adverse event for the Unknown COVID vaccine was headache (35%) for first dose and three participants reported fever as their adverse event for their second dose. The most frequently reported adverse event for the Flu vaccine was injection site complication (32%). At this age group, the most common adverse events across the five-vaccine cohort were fever, injection site complication and headache (Table 2).

For the +65-year age group, the Moderna cohort had the largest number of reports compared to the other four cohorts. The most frequently reported adverse events for the Pfizer vaccine were fever (17%) and nonspecific musculoskeletal pain (17%) for the first dose, and dyspnea NOS (15%) for the second dose. The most frequently reported adverse events for the Moderna vaccine were injection site complication (29%) for the first dose and fever (19%) for the second dose. The most frequently reported adverse event for Janssen vaccine was Fever (20%) for the first dose. The most frequently reported adverse event for the Unknown COVID vaccine were fever (26%) for the first dose and fatigue (33%) for the second dose. The most frequently reported adverse event for the Flu vaccine was injection site complications (33%). At this age group, the most common adverse events across the five-vaccine cohort were fever, injection site complication, nonspecific musculoskeletal pain, and fatigue (Table 2). Supplemental tables list risk percentages for additional adverse events reported in the VAERS (**Supplemental Table S3**).

### 3.2 Comparison of adverse events for COVID-19 and Flu Vaccines

Among the common adverse events between the Flu and COVID-19 vaccines, the Relative Risk along with 95% Confidence Intervals were calculated to facilitate comparisons of adverse events between groups. For the age group of 12-15, persons receiving first dose of Pfizer vaccine were 0.64 times more likely to experience pallor (95% CI: 0.44 to 0.95), 0.55 times more likely to experience hypotension (95% CI:0.45 to 0.68), 0.49 times more likely to experience visual changes (95% CI: 0.32 to 0.73) than a person receiving Flu vaccine. Persons receiving second dose of Pfizer vaccine were 0.37 times more likely to experience central neuropathy compared to those receiving the Flu vaccine (95% CI: 0.22 to 0.63) (Table 3).

**Table 3:**
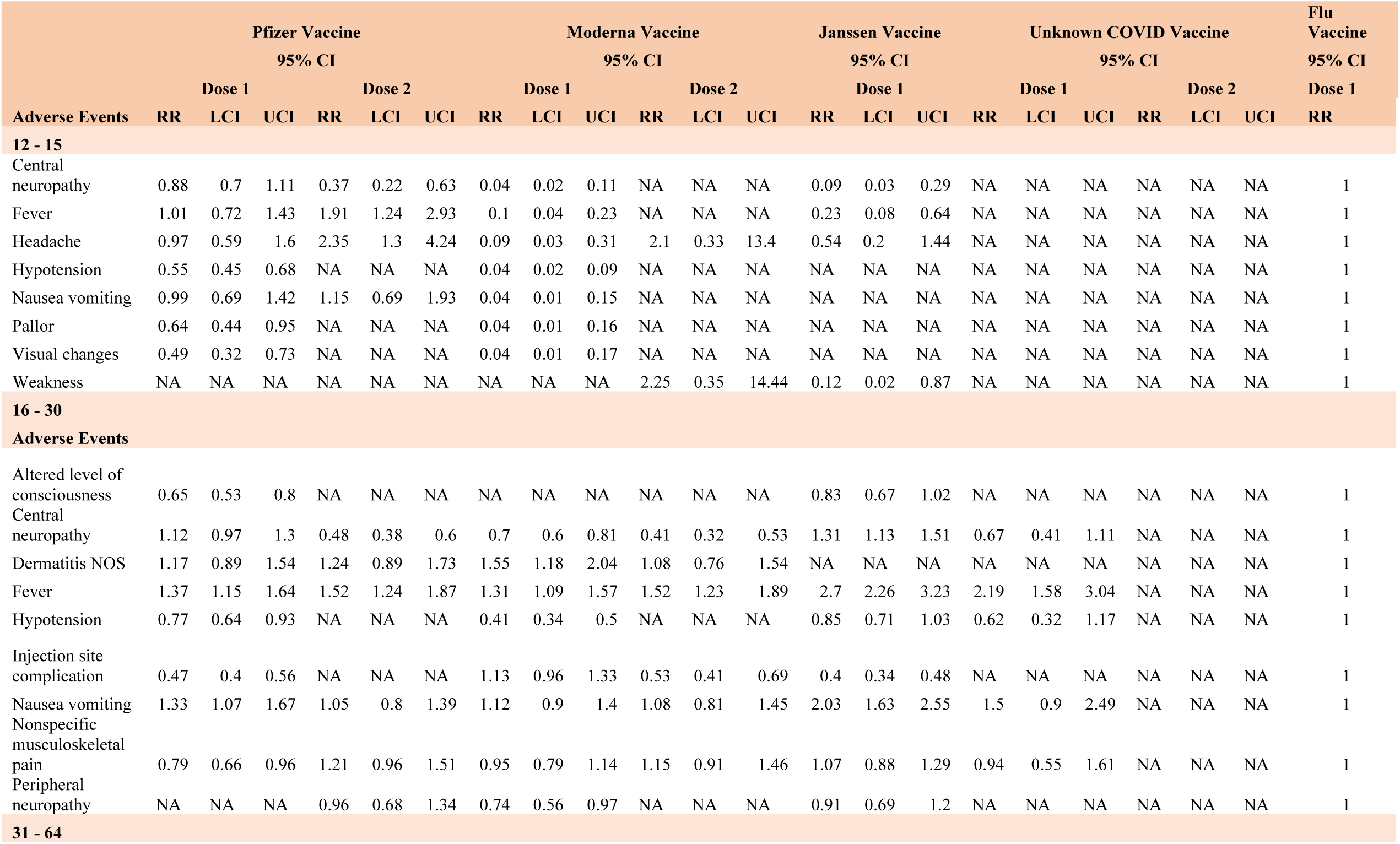
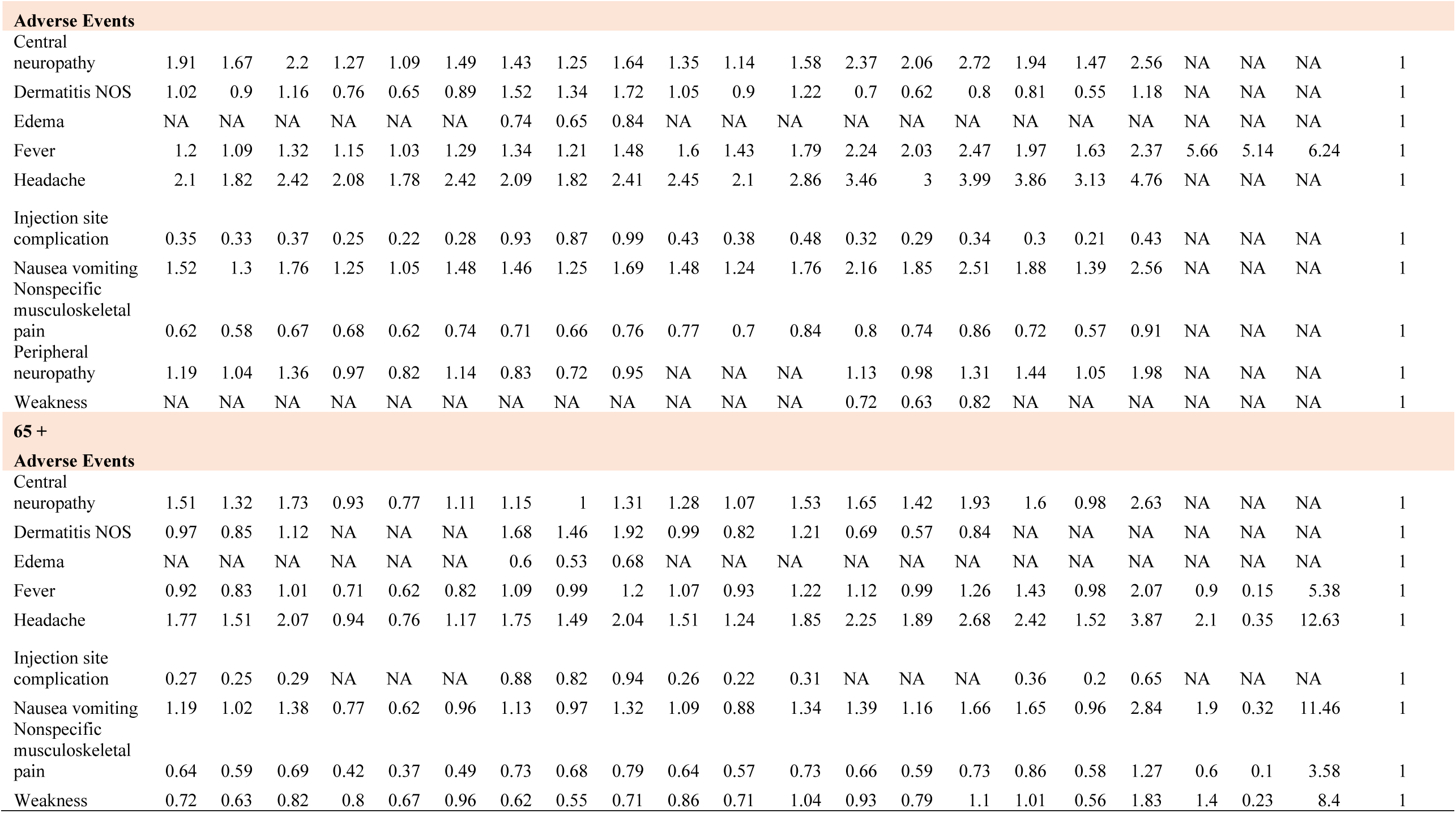
Relative risks and associated confidence intervals for most frequently reported adverse events.

Persons from the 12 to 15-year age group who received first dose of the Moderna vaccine were 0.1 times more likely to experience fever (95% CI: 0.04 to 0.23), 0.09 times more likely to experience headache (95% CI: 0.03 to 0.31), 0.04 times more likely to experience central neuropathy (95% CI: 0.02 to 0.11), 0.04 times more likely to experience hypotension (95% CI: 0.02 to 0.09), 0.04 times more likely to experience nausea vomiting (95% CI: 0.01 to 0.15), 0.04 times more likely to experience pallor (95% CI: 0.01 to 0.16), and 0.04 times more likely to experience visual changes (95% CI: 0.01 to 0.17) than person receiving the Flu vaccine. Relative risks reported for persons receiving second dose of Moderna vaccine were not statistically significant (Table 3).

Persons receiving first dose of Janssen vaccine were 0.23 times more likely to experience fever (95% CI: 0.08 to 0.64), 0.12 times more likely to experience weakness (95% CI: 0.02 to 0.87), and 0.09 times more likely to experience central neuropathy (95% CI: 0.03 to 0.29) than persons receiving the Flu vaccine (Table 3).

For the age group of 16-30, persons receiving first dose of the Pfizer vaccine were 1.37 times more likely to experience fever (95% CI:1.15 to 1.64), 0.79 times more likely to experience nonspecific musculoskeletal pain (95% CI: 0.66 to 0.96), 0.77 times more likely to experience hypotension (95% CI: 0.64 to 0.93), 0.65 time more likely to experience altered level of consciousness (95% CI: 0.53 to 0.8), and 0.47 times more likely to experience injection site complications (95% CI: 0.4 to 0.56) than person receiving Flu vaccine. Persons receiving second dose of the Pfizer vaccine were 1.52 times more likely to experience fever (95% CI:1.24 to 1.87) and 0.48 times more likely to experience central neuropathy (95% CI: 0.38 to 0.6) than person receiving the Flu vaccine (Table 3).

Persons receiving first dose of the Moderna vaccine were 0.74 times more likely to experience peripheral neuropathy (95% CI: 0.56 to 0.97), 0.7 times more likely to experience central neuropathy (95% CI: 0.6 to 0.81), and 0.4 times more likely to experience hypotension (95% CI: 0.34 to 0.5) than person receiving Flu vaccine. Persons receiving second dose of the Moderna vaccine were 1.52 times more likely to experience fever (95% CI: 1.23 to 1.89), 0.53 times more likely to experience injection site complications (95% CI: 0.41 to 0.69), and 0.41 times more likely to experience central neuropathy (95% CI: 0.32 to 0.53) than person receiving Flu vaccine (Table 3).

Persons receiving the Janssen vaccine were 2.03 times more likely to experience Nausea vomiting (95% CI: 1.63 to 2.55), 1.31 times more likely to experience central neuropathy (95% CI: 1.13 to 1.51), and 0.4 times more likely to experience injection site complications (95% CI: 0.34 to 0.48) than person receiving Flu vaccine (Table 3). Persons receiving the Unknown COVID vaccine were 2.19 times more likely to experience fever (95% CI: 1.58 to 3.04) compared to persons receiving the Flu vaccine.

For the 31 to 64-year age group, persons receiving first dose of the Pfizer vaccine were 2.1 times more likely to experience headache (95% CI: 1.82 to 2.42), 1.91 times more likely to experience central neuropathy (95% CI: 1.67 to 2.2), 1.52 times more likely to experience nausea vomiting (95% CI: 1.3 to 1.76), 1.2 times more likely to experience fever (95% CI: 1.09 to 1.32), 1.19 times more likely to experience peripheral neuropathy (95% CI: 1.04 to 1.36), 0.62 times more likely to experience nonspecific musculoskeletal pain (95% CI: 0.58 to 0.67), and 0.35 times more likely to experience injection site complications (95% CI: 0.33 to 0.37) compared to reports of those taking the Flu vaccine (Table 3).

Persons receiving second dose of the Pfizer vaccine were 2.08 times more likely to experience headache (95% CI: 1.78 to 2.42), 1.27 times more likely to experience central neuropathy (95% CI: 1.09 to 1.49), 1.25 times more likely to experience nausea vomiting (95% CI: 1.05 to 1.48), 1.15 times more likely to experience fever (95% CI: 1.03 to 1.29), 0.76 times more likely to experience dermatitis NOS (95% CI: 0.65 to 0.89), 0.68 times more likely to experience nonspecific musculoskeletal pain (95% CI: 0.62 to 0.74), and 0.25 times more likely to experience injection site complications (95% CI: 0.22 to 0.28) than person receiving the Flu vaccine (Table 3).

Persons receiving first dose of the Moderna vaccine were 2.09 times more likely to experience headache(95% CI: 1.82 to 2.41), 1.52 times more likely to experience dermatitis NOS (95% CI: 1.34 to 1.72), 1.46 times more likely to experience nausea vomiting (95% CI: 1.25 to 1.69),1.43 times more likely to experience central neuropathy (95% CI: 1.25 to 1.64), 1.34 times more likely to experience fever (95% CI: 1.21 to 1.48), 0.93 times more likely to experience injection site complications (95% CI: 0.87 to 0.99), 0.83 times more likely to experience peripheral neuropathy (95% CI: 0.72 to 0.95), 0.74 times more likely to experience edema (95% CI: 0.65 to 0.84), and 0.71 times more likely to experience nonspecific musculoskeletal pain (95% CI: 0.66 to 0.76) than person receiving the Flu vaccine (Table 3).

Persons receiving second dose of the Moderna vaccine were 2.45 times more likely to get headache (95% CI: 2.1 to 2.86), 1.6 times more likely to experience fever (95% CI: 1.43 to 1.79), 1.48 times more likely to get nausea vomiting (95% CI: 1.24 to 1.76), 1.35 times more likely to get central neuropathy (95% CI: 1.14 to 1.58), 0.77 times more likely to experience nonspecific musculoskeletal pain (95% CI: 0.7 to 0.84), and 0.43 times more likely to experience injection site complications (95% CI: 0.38 to 0.48) than person receiving the Flu vaccine (Table 3).

Persons receiving the first dose of Janssen vaccine were 3.46 times more likely to get headache (95% CI: 3 to 3.99), 2.37 times more likely to get central neuropathy (95% CI: 2.06 to 2.76), 2.24 times more likely to get fever (95% CI: 2.03 to 2.47), 2.16 times more likely to get nausea vomiting (95% CI: 1.85 to 2.51), 0.8 times more likely to experience nonspecific musculoskeletal pain (95% CI: 0.74 to 0.86), 0.72 times more likely to experience weakness (95% CI: 0.63 to 0.82), 0.7 times more likely to experience dermatitis NOS (95% CI: 0.62 to 0.8), and 0.32 times more likely to experience injection site complications (95% CI: 0.29 to 0.34) than a person receiving the Flu vaccine (Table 3).

Persons receiving the first dose of the Unknown COVID vaccine were 3.86 times more likely to experience headache (95% CI: 3.13 to 4.76), 1.97 times more likely to experience fever (95% CI: 1.63 to 2.37), 1.94 times more likely to experience central neuropathy (95% CI: 1.47 to 2.56), 1.88 times more likely to experience nausea vomiting (95% CI: 1.39 to 2.56), 1.44 times more likely to experience peripheral neuropathy (95% CI: 1.05 to 1.98), 0.72 times more likely to experience nonspecific musculoskeletal pain (95% CI: 0.57 to 0.91), and 0.3 times more likely to experience injection site complications (95% CI: 0.21 to 0.43) than person receiving the Flu vaccine (Table 3).

For 65+ age group, persons receiving first dose of the Pfizer vaccine were 1.77 times more likely to experience headache (95% CI: 1.51 to 2.07), 1.51 times more likely to experience central neuropathy (95% CI: 1.3 to 1.73), 1.19 times more likely to experience nausea vomiting (95% CI: 1.02 to 1.38), 0.72 times more likely to experience weakness (95% CI: 0.63 to 0.82), 0.64 times more likely to experience nonspecific musculoskeletal pain (95% CI: 0.59 to 0.69), and 0.27 times more likely to experience injection site complication (95% CI: 0.25 to 0.29) than person receiving the Flu vaccine. Persons receiving second dose of Pfizer vaccine were 0.8 times more likely to experience weakness (95% CI: 0.67 to 0.96), 0.77 times more likely to experience nausea and vomiting (95% CI: 0.62 to 0.96), 0.71 times more likely to experience fever (95% CI: 0.62 to 0.82), and 0.42 times more likely to experience nonspecific musculoskeletal pain (95% CI: 0.37 to 0.49) than a person receiving the Flu vaccine (Table 3).

Persons receiving the first dose of the Moderna vaccine were 1.75 times more likely to experience headache (95% CI: 1.49 to 2.04), 1.68 times more likely to experience dermatitis NOS (95% CI: 1.46 to 1.92), 0.88 times more likely to experience injection site complications (95% CI: 0.82 to 0.94), 0.73 times more likely to experience nonspecific musculoskeletal pain (95% CI: 0.68 to 0.79), 0.62 times more likely to experience weakness (95% CI: 0.55 to 0.71), and 0.6 times more likely to experience edema (95% CI: 0.53 to 0.68) than a person receiving the Flu vaccine (Table 3).

Persons receiving second dose of the Moderna vaccine were 1.51 times more likely to experience headache (95% CI: 1.24 to 1.85), 1.28 times more likely to experience central neuropathy (95% CI: 1.07 to 1.53), 0.64 times more likely to experience nonspecific musculoskeletal pain (95% CI: 0.57 to 0.73), and 0.26 times more likely to experience injection site complications (95% CI: 0.22 to 0.31) than a person receiving the Flu vaccine.

Persons receiving Janssen vaccine were 2.25 times more likely to experience headache (95% CI: 1.89 to 2.68), 1.65 times more likely to experience central neuropathy (95% CI: 1.42 to 1.93), 1.39 times more likely to experience nausea vomiting (95% CI: 1.16 to 1.66), 0.69 times more likely to experience dermatitis NOS (95% CI: 0.57 to 0.84), and 0.66 times more likely to experience nonspecific musculoskeletal pain (95% CI: 0.59 to 0.73) than a person receiving the Flu vaccine (Table 3).

Persons receiving the Unknown COVID vaccine were 2.42 times more likely to experience headache (95% CI: 1.52 to 3.87) and 0.36 times more likely to experience injection site complications (95% CI: 0.2 to 0.65) than person receiving Flu vaccine (Table 3).

Apart from the frequently reported adverse events (Table 3), some adverse events were worth mentioning due to the growing concerns in the media, such as Guillain Barre Syndrome[26], and gynecologic changes[27]. Since these concerns were specific to COVID-19 vaccine and rarely reported as adverse events for the Flu vaccines, relative risk statistics were not calculated. Such rare events have been identified in our study and have been listed in the Supplemental table (**Supplemental table S3**).

## 4. Discussions

As mentioned previously, the interpretation of the results is subjected to limitations of passive surveillance system. Moderna had the highest number of reports in VAERS among all cohorts. Most of the reports were from the 31-64 age group which is likely due to the initial authorization of vaccine for the 31-64 age group. The proportion of reports were equally divided among having recovered and not recovered from the adverse event across all cohorts. The proportion of death across COVID vaccines cohorts were comparable and may indicate the high fatality rate of the disease rather than the vaccines especially given that the COVID-19 vaccines were recently introduced and those with severe symptoms had earlier access to them than the general population.

Across the five cohorts and age groups, common adverse events could be summarized as: central neuropathy, fever, headache, injection site, nonspecific musculoskeletal pain, chest pain. Except for chest pain, most of the adverse events were already identified in randomized controlled clinical trials of COVID-19 vaccines. Our study provides an age-wise distribution of adverse events as reported in the VAERS database (Table 2). It is hoped that this information will assist some in overcoming their vaccine hesitancy due to safety concerns.

Investigating rare side effects of any vaccine is a difficult task[28] due to the rarity of some side-effects, the longer period of manifestation for some side-effects, and lack of evidence for causation Observational studies like that reported here is often is a good starting point to higher quality research. A robust surveillance system (usually a combination of active and passive surveillance systems) is key in documenting rare adverse events and generating hypotheses for randomized controlled trials or case-control studies.

It is important to interpret these results considering their limitations. The lack of availability of time to each adverse event reported prevented time-to-event analysis such as survival analysis (i.e., Kaplan-Meier Survival curves and Cox proportional hazard regression). Potential selection bias could apply to adverse event reporting in cases of COVID-19 vaccinations compared to adverse events reported in cases of Flu vaccinations. Finally, reporting of adverse events to VAERS does not imply that the adverse event is caused by the immunization event.

## 5. Conclusions

Based on the available data and results, it appears that there were some common adverse events between Flu vaccines and COVID-19 vaccines. These identified common adverse events warrant further investigations based on the relative risk and 95% CI.

Continued vaccine safety monitoring and ongoing advocacy targeting public for reporting of any adverse events associated with available COVID-19 vaccines to the VAERS will help researchers to identify adverse events related with vaccines and generate hypotheses for further investigations. The current study can be used by laypersons to inform decision-making about COVID-19 and Flu vaccinations to overcome one aspect of vaccine hesitancy.

## Data Availability

Data is publicly available at VAERS.

## Acknowledgments

We would like to acknowledge the help of Ms. Marta Shore, MS Lecturer of Division of Biostatistics, School of Public Health, University of Minnesota for her expert review of the methods and Mr. Ariel Roane, Institute for Health Informatics, University of Minnesota for his review and editing of the manuscript.

