## Supplemental Tables for "Comparison of adverse events between COVID-19 and Flu vaccines"

| Number | VAERS Adverse event | Re-grouped Adverse Events |
|---|---|---|
| 1 | Abdominal discomfort | Abdominal pain/discomfort |
| 2 | Abdominal distension | Abdominal pain/discomfort |
| 3 | Abdominal pain | Abdominal pain/discomfort |
| 4 | Abdominal pain upper | Abdominal pain/discomfort |
| 5 | Dyspepsia | Abdominal pain/discomfort |
| 6 | Gastrooesophageal reflux disease | Abdominal pain/discomfort |
| 7 | Pelvic pain | Abdominal pain/discomfort |
| 8 | Alopecia | Alopecia |
| 9 | Altered state of consciousness | Altered level of consciousness |
| 10 | Consciousness fluctuating | Altered level of consciousness |
| 11 | Delirium | Altered level of consciousness |
| 12 | Depressed level of consciousness | Altered level of consciousness |
| 13 | Disorientation | Altered level of consciousness |
| 14 | Disturbance in attention | Altered level of consciousness |
| 15 | Dreamy state | Altered level of consciousness |
| 16 | Incoherent | Altered level of consciousness |
| 17 | Loss of consciousness | Altered level of consciousness |
| 18 | Mental impairment | Altered level of consciousness |
| 19 | Mental status changes | Altered level of consciousness |
| 20 | Respiratory rate decreased | Altered level of consciousness |
| 21 | Unresponsive to stimuli | Altered level of consciousness |
| 22 | Hyperaesthesia | Altered sensation |
| 23 | Anaemia | Anaemia |
| 24 | Anaphylactic reaction | Anaphylaxis |
| 25 | Anaphylactic shock | Anaphylaxis |
| 26 | Decreased appetite | Anorexia |
| 27 | Feeding disorder | Anorexia |
| 28 | Joint noise | Arthropathy |
| 29 | Asthma | Asthma exacerbation |
| 30 | Autonomic nervous system imbalance | Autonomic neuropathy |
| 31 | Axillary pain | Axillary pain |
| 32 | Beta haemolytic streptococcal infection | Bacterial infection NOS |
| 33 | Streptococcus test positive | Bacterial infection NOS |
| 34 | Urinary tract infection | Bacterial infection NOS |
| 35 | Urine odour abnormal | Bacterial infection NOS |
| 36 | Haemorrhage | Bleeding NOS |
| 37 | Constipation | Bowel pattern changes |
| 38 | Diarrhoea | Bowel pattern changes |
| 39 | Flatulence | Bowel pattern changes |
| 40 | Bradycardia | Bradycardia |
| 41 | Heart rate decreased | Bradycardia |
| 42 | Breast pain | Breast pain |
| 43 | Cardiac arrest | Cardiorespiratory arrest |
| 44 | Cardio-respiratory arrest | Cardiorespiratory arrest |
| 45 | Encephalitis | Central neuritis |
| 46 | Encephalitis autoimmune | Central neuritis |
| 47 | Encephalomyelitis | Central neuritis |
| 48 | Acute disseminated encephalomyelitis | Central neuropathy |
| 49 | Coordination abnormal | Central neuropathy |
| 50 | Dizziness | Central neuropathy |
| 51 | Dizziness postural | Central neuropathy |
| 52 | Dyskinesia | Central neuropathy |
| 53 | Dysstasia | Central neuropathy |
| 54 | Fall | Central neuropathy |
| 55 | Gait disturbance | Central neuropathy |
| 56 | Gait inability | Central neuropathy |

| | | |
|---|---|---|
| 57 | Hemiparesis | Central neuropathy |
| 58 | Myelitis transverse | Central neuropathy |
| 59 | Nervous system disorder | Central neuropathy |
| 60 | Paralysis | Central neuropathy |
| 61 | Sensory disturbance | Central neuropathy |
| 62 | Sensory loss | Central neuropathy |
| 63 | Tic | Central neuropathy |
| 64 | Tremor | Central neuropathy |
| 65 | Vertigo | Central neuropathy |
| 66 | Cerebral haemorrhage | Cerebral hemorrhage |
| 67 | Subdural haematoma | Cerebral hemorrhage |
| 68 | Cerebral infarction | Cerebral thrombosis/infarction |
| 69 | Cerebral thrombosis | Cerebral thrombosis/infarction |
| 70 | Cerebrovascular accident | Cerebral thrombosis/infarction |
| 71 | Embolic stroke | Cerebral thrombosis/infarction |
| 72 | Ischaemic stroke | Cerebral thrombosis/infarction |
| 73 | Transient ischaemic attack | Cerebral thrombosis/infarction |
| 74 | Chest discomfort | Chest pain |
| 75 | Chest pain | Chest pain |
| 76 | Choking | Choking |
| 77 | Cognitive disorder | Cognitive disorder |
| 78 | Confusional state | Cognitive disorder |
| 79 | Thinking abnormal | Cognitive disorder |
| 80 | Cardiac failure congestive | Congestive heart failure |
| 81 | Chronic obstructive pulmonary disease | COPD exacerbation |
| 82 | Cough | Cough |
| 83 | Productive cough | Cough |
| 84 | Death | Death |
| 85 | Deep vein thrombosis | Deep vein thrombosis |
| 86 | Dehydration | Dehydration |
| 87 | Arrested labour | Delivery complication |
| 88 | Dermatitis bullous | Dermatitis NOS |
| 89 | Eczema | Dermatitis NOS |
| 90 | Pruritus | Dermatitis NOS |
| 91 | Rash | Dermatitis NOS |
| 92 | Rash erythematous | Dermatitis NOS |
| 93 | Rash macular | Dermatitis NOS |
| 94 | Rash papular | Dermatitis NOS |
| 95 | Rash pruritic | Dermatitis NOS |
| 96 | Rash vesicular | Dermatitis NOS |
| 97 | Sensitive skin | Dermatitis NOS |
| 98 | Skin exfoliation | Dermatitis NOS |
| 99 | Skin irritation | Dermatitis NOS |
| 100 | Skin reaction | Dermatitis NOS |
| 101 | Dry skin | Dermatitis NOS |
| 102 | Balance disorder | Dizziness |
| 103 | Drooling | Drooling |
| 104 | Dry mouth | Dry mouth |
| 105 | Dysphagia | Dysphagia |
| 106 | Hypophagia | Dysphagia |
| 107 | Atelectasis | Dyspnea NOS |
| 108 | Dyspnoea | Dyspnea NOS |
| 109 | Dyspnoea exertional | Dyspnea NOS |
| 110 | Hyperventilation | Dyspnea NOS |
| 111 | Hypopnoea | Dyspnea NOS |
| 112 | Hypoxia | Dyspnea NOS |
| 113 | Oxygen saturation decreased | Dyspnea NOS |

| | | |
|---|---|---|
| 114 | Pleural effusion | Dyspnea NOS |
| 115 | Pulmonary oedema | Dyspnea NOS |
| 116 | Respiratory rate increased | Dyspnea NOS |
| 117 | Respiratory tract congestion | Dyspnea NOS |
| 118 | Tachypnoea | Dyspnea NOS |
| 119 | Wheezing | Dyspnea NOS |
| 120 | Ear discomfort | Ear pain/discomfort |
| 121 | Ear pain | Ear pain/discomfort |
| 122 | Oedema | Edema |
| 123 | Oedema peripheral | Edema |
| 124 | Peripheral swelling | Edema |
| 125 | Skin swelling | Edema |
| 126 | Skin tightness | Edema |
| 127 | Swelling | Edema |
| 128 | Swelling face | Edema |
| 129 | Hot flush | Endocrine disorder |
| 130 | Erythema | Erythema NOS |
| 131 | Dry eye | Eye irritation |
| 132 | Excessive eye blinking | Eye irritation |
| 133 | Eye discharge | Eye irritation |
| 134 | Eye disorder | Eye irritation |
| 135 | Eye irritation | Eye irritation |
| 136 | Eye pain | Eye irritation |
| 137 | Eye pruritus | Eye irritation |
| 138 | Eye swelling | Eye irritation |
| 139 | Lacrimation increased | Eye irritation |
| 140 | Ocular hyperaemia | Eye irritation |
| 141 | Periorbital swelling | Eye irritation |
| 142 | Swelling of eyelid | Eye irritation |
| 143 | Bell's palsy | Facial neuropathy |
| 144 | Facial pain | Facial neuropathy |
| 145 | Facial paralysis | Facial neuropathy |
| 146 | Facial paresis | Facial neuropathy |
| 147 | Bedridden | Failure to thrive |
| 148 | General physical health deterioration | Failure to thrive |
| 149 | Impaired work ability | Failure to thrive |
| 150 | Lethargy | Failure to thrive |
| 151 | Loss of personal independence in daily activities | Failure to thrive |
| 152 | Malnutrition | Failure to thrive |
| 153 | Staring | Failure to thrive |
| 154 | Weight decreased | Failure to thrive |
| 155 | Fatigue | Fatigue |
| 156 | Somnolence | Fatigue |
| 157 | Abortion spontaneous | Fetal death |
| 158 | Foetal death | Fetal death |
| 159 | Body temperature increased | Fever |
| 160 | Chills | Fever |
| 161 | Cold sweat | Fever |
| 162 | Feeling cold | Fever |
| 163 | Feeling hot | Fever |
| 164 | Feeling of body temperature change | Fever |
| 165 | Hyperhidrosis | Fever |
| 166 | Night sweats | Fever |
| 167 | Peripheral coldness | Fever |
| 168 | Pyrexia | Fever |
| 169 | Flushing | Flushing |
| 170 | Bursitis | Focal musculoskeletal pain |

| | | |
|---|---|---|
| 171 | Pain in jaw | Focal musculoskeletal pain |
| 172 | Rotator cuff syndrome | Focal musculoskeletal pain |
| 173 | Tendon disorder | Focal musculoskeletal pain |
| 174 | Tendonitis | Focal musculoskeletal pain |
| 175 | Blister | Focal skin abnormality |
| 176 | Gastrointestinal haemorrhage | Gastrointestinal bleeding |
| 177 | Mast cell activation syndrome | Generalized allergic response |
| 178 | Groin pain | Groin pain |
| 179 | Guillain-Barre syndrome | Guillain-Barre syndrome |
| 180 | Dysmenorrhoea | Gynecologic changes |
| 181 | Heavy menstrual bleeding | Gynecologic changes |
| 182 | Menstrual disorder | Gynecologic changes |
| 183 | Menstruation irregular | Gynecologic changes |
| 184 | Vaginal haemorrhage | Gynecologic changes |
| 185 | Concussion | Head trauma |
| 186 | Head discomfort | Headache |
| 187 | Headache | Headache |
| 188 | Migraine | Headache |
| 189 | Deafness | Hearing changes |
| 190 | Deafness unilateral | Hearing changes |
| 191 | Hyperacusis | Hearing changes |
| 192 | Hypoacusis | Hearing changes |
| 193 | Tinnitus | Hearing changes |
| 194 | Blood urine present | Hematuria |
| 195 | Haematuria | Hematuria |
| 196 | Autoimmune hepatitis | Hepatitis |
| 197 | Hepatitis | Hepatitis |
| 198 | Hepatitis acute | Hepatitis |
| 199 | Jaundice | Hepatitis |
| 200 | Liver injury | Hepatitis |
| 201 | Ocular icterus | Hepatitis |
| 202 | Hypertension | Hypertension |
| 203 | Hypotension | Hypotension |
| 204 | Orthostatic hypotension | Hypotension |
| 205 | Postural orthostatic tachycardia syndrome | Hypotension |
| 206 | Presyncope | Hypotension |
| 207 | Syncope | Hypotension |
| 208 | Body temperature decreased | Hypothermia |
| 209 | Temperature regulation disorder | Hypothermia |
| 210 | Thirst | Hypovolemia |
| 211 | Contusion | Injection site complication |
| 212 | Extensive swelling of vaccinated limb | Injection site complication |
| 213 | Injected limb mobility decreased | Injection site complication |
| 214 | Injection site bruising | Injection site complication |
| 215 | Injection site cellulitis | Injection site complication |
| 216 | Injection site discolouration | Injection site complication |
| 217 | Injection site discomfort | Injection site complication |
| 218 | Injection site erythema | Injection site complication |
| 219 | Injection site extravasation | Injection site complication |
| 220 | Injection site haemorrhage | Injection site complication |
| 221 | Injection site hypoaesthesia | Injection site complication |
| 222 | Injection site induration | Injection site complication |
| 223 | Injection site infection | Injection site complication |
| 224 | Injection site inflammation | Injection site complication |
| 225 | Injection site irritation | Injection site complication |
| 226 | Injection site mass | Injection site complication |
| 227 | Injection site nodule | Injection site complication |

| | | |
|---|---|---|
| 228 | Injection site oedema | Injection site complication |
| 229 | Injection site pain | Injection site complication |
| 230 | Injection site papule | Injection site complication |
| 231 | Injection site paraesthesia | Injection site complication |
| 232 | Injection site pruritus | Injection site complication |
| 233 | Injection site rash | Injection site complication |
| 234 | Injection site reaction | Injection site complication |
| 235 | Injection site swelling | Injection site complication |
| 236 | Injection site urticaria | Injection site complication |
| 237 | Injection site vesicles | Injection site complication |
| 238 | Injection site warmth | Injection site complication |
| 239 | Local reaction | Injection site complication |
| 240 | Localised infection | Injection site complication |
| 241 | Sensation of foreign body | Injection site complication |
| 242 | Shoulder injury related to vaccine administration | Injection site complication |
| 243 | Thrombophlebitis superficial | Injection site complication |
| 244 | Vaccination site erythema | Injection site complication |
| 245 | Vaccination site pain | Injection site complication |
| 246 | Vaccination site pruritus | Injection site complication |
| 247 | Vaccination site rash | Injection site complication |
| 248 | Vaccination site reaction | Injection site complication |
| 249 | Vaccination site swelling | Injection site complication |
| 250 | Vaccination site warmth | Injection site complication |
| 251 | Arrhythmia | Irregular heart rate |
| 252 | Atrial fibrillation | Irregular heart rate |
| 253 | Heart rate irregular | Irregular heart rate |
| 254 | Lymph node pain | Lymphadenopathy |
| 255 | Lymphadenopathy | Lymphadenopathy |
| 256 | Discomfort | Malaise |
| 257 | Feeling abnormal | Malaise |
| 258 | Illness | Malaise |
| 259 | Malaise | Malaise |
| 260 | Amnesia | Memory disorder |
| 261 | Memory impairment | Memory disorder |
| 262 | Acute myocardial infarction | Myocardial infarction |
| 263 | Myocardial infarction | Myocardial infarction |
| 264 | Myocarditis | Myocarditis |
| 265 | Nausea | Nausea/vomiting |
| 266 | Retching | Nausea/vomiting |
| 267 | Vomiting | Nausea/vomiting |
| 268 | Vomiting projectile | Nausea/vomiting |
| 269 | Back pain | Neck/back pain |
| 270 | Neck pain | Neck/back pain |
| 271 | Spinal pain | Neck/back pain |
| 272 | Acute flaccid myelitis | Neuromuscular disorder |
| 273 | Burning sensation | Neuropathy NOS |
| 274 | Arthralgia | Nonspecific musculoskeletal pain |
| 275 | Arthritis | Nonspecific musculoskeletal pain |
| 276 | Bone pain | Nonspecific musculoskeletal pain |
| 277 | Joint range of motion decreased | Nonspecific musculoskeletal pain |
| 278 | Joint stiffness | Nonspecific musculoskeletal pain |
| 279 | Joint swelling | Nonspecific musculoskeletal pain |
| 280 | Limb discomfort | Nonspecific musculoskeletal pain |
| 281 | Muscle strain | Nonspecific musculoskeletal pain |
| 282 | Muscle tightness | Nonspecific musculoskeletal pain |
| 283 | Musculoskeletal chest pain | Nonspecific musculoskeletal pain |
| 284 | Musculoskeletal discomfort | Nonspecific musculoskeletal pain |

| | | |
|---|---|---|
| 285 | Musculoskeletal pain | Nonspecific musculoskeletal pain |
| 286 | Musculoskeletal stiffness | Nonspecific musculoskeletal pain |
| 287 | Myalgia | Nonspecific musculoskeletal pain |
| 288 | Pain in extremity | Nonspecific musculoskeletal pain |
| 289 | Periarthritis | Nonspecific musculoskeletal pain |
| 290 | Epistaxis | Nosebleed |
| 291 | Herpes simplex | Other viral infection |
| 292 | Herpes zoster | Other viral infection |
| 293 | Oral herpes | Other viral infection |
| 294 | Overdose | Overdose |
| 295 | Cyanosis | Pallor |
| 296 | Pallor | Pallor |
| 297 | Pericarditis | Pericarditis |
| 298 | Hypoaesthesia oral | Perioral inflammation |
| 299 | Lip erythema | Perioral inflammation |
| 300 | Lip swelling | Perioral inflammation |
| 301 | Mouth swelling | Perioral inflammation |
| 302 | Oral pruritus | Perioral inflammation |
| 303 | Paraesthesia oral | Perioral inflammation |
| 304 | Hypoaesthesia | Peripheral neuropathy |
| 305 | Muscle contractions involuntary | Peripheral neuropathy |
| 306 | Muscle rigidity | Peripheral neuropathy |
| 307 | Muscle spasms | Peripheral neuropathy |
| 308 | Muscle twitching | Peripheral neuropathy |
| 309 | Neuralgia | Peripheral neuropathy |
| 310 | Neuropathy peripheral | Peripheral neuropathy |
| 311 | Opisthotonus | Peripheral neuropathy |
| 312 | Paraesthesia | Peripheral neuropathy |
| 313 | Skin burning sensation | Peripheral neuropathy |
| 314 | Monoplegia | Peripheral neuropathy |
| 315 | Peripheral embolism | Peripheral thrombosis |
| 316 | Dry throat | Pharyngitis |
| 317 | Nasopharyngitis | Pharyngitis |
| 318 | Oropharyngeal discomfort | Pharyngitis |
| 319 | Oropharyngeal pain | Pharyngitis |
| 320 | Pharyngeal paraesthesia | Pharyngitis |
| 321 | Pharyngeal swelling | Pharyngitis |
| 322 | Throat clearing | Pharyngitis |
| 323 | Throat irritation | Pharyngitis |
| 324 | Tonsillar hypertrophy | Pharyngitis |
| 325 | Vocal cord disorder | Pharyngitis |
| 326 | Lung infiltration | Pneumonia |
| 327 | Lung opacity | Pneumonia |
| 328 | Pneumonia | Pneumonia |
| 329 | Amniotic cavity infection | Pregnancy complication |
| 330 | Caesarean section | Pregnancy complication |
| 331 | Complication of pregnancy | Pregnancy complication |
| 332 | Foetal disorder | Pregnancy complication |
| 333 | Foetal growth restriction | Pregnancy complication |
| 334 | Foetal heart rate abnormal | Pregnancy complication |
| 335 | Gestational diabetes | Pregnancy complication |
| 336 | Gestational hypertension | Pregnancy complication |
| 337 | Neonatal disorder | Pregnancy complication |
| 338 | Pre-eclampsia | Pregnancy complication |
| 339 | Premature delivery | Pregnancy complication |
| 340 | Premature labour | Pregnancy complication |
| 341 | Prolonged labour | Pregnancy complication |

| | | |
|---|---|---|
| 342 | Aggression | Psychiatric amplification |
| 343 | Agitation | Psychiatric amplification |
| 344 | Anxiety | Psychiatric amplification |
| 345 | Conversion disorder | Psychiatric amplification |
| 346 | Emotional distress | Psychiatric amplification |
| 347 | Hallucination | Psychiatric amplification |
| 348 | Intrusive thoughts | Psychiatric amplification |
| 349 | Irritability | Psychiatric amplification |
| 350 | Nervousness | Psychiatric amplification |
| 351 | Obsessive-compulsive disorder | Psychiatric amplification |
| 352 | Panic attack | Psychiatric amplification |
| 353 | Restlessness | Psychiatric amplification |
| 354 | Social avoidant behaviour | Psychiatric amplification |
| 355 | Crying | Psychiatric depression |
| 356 | Depression | Psychiatric depression |
| 357 | Intentional self-injury | Psychiatric depression |
| 358 | Suicidal behaviour | Psychiatric depression |
| 359 | Neuropsychiatric syndrome | Psychiatric disorder |
| 360 | Paediatric autoimmune neuropsychiatric disorders associated with streptococcal infection | Psychiatric disorder |
| 361 | Pulmonary embolism | Pulmonary embolism |
| 362 | Pulmonary thrombosis | Pulmonary embolism |
| 363 | Butterfly rash | Rash |
| 364 | Urticaria | Rash |
| 365 | Raynaud's phenomenon | Raynaud's phenomenon |
| 366 | Acute kidney injury | Renal failure |
| 367 | Renal failure | Renal failure |
| 368 | Respiratory arrest | Respiratory failure |
| 369 | Respiratory distress | Respiratory failure |
| 370 | Respiratory failure | Respiratory failure |
| 371 | Acute respiratory failure | Respiratory failure |
| 372 | Nasal congestion | Rhinitis |
| 373 | Paranasal sinus discomfort | Rhinitis |
| 374 | Rhinorrhoea | Rhinitis |
| 375 | Sinusitis | Rhinitis |
| 376 | Sneezing | Rhinitis |
| 377 | Generalised tonic-clonic seizure | Seizure |
| 378 | Postictal state | Seizure |
| 379 | Seizure | Seizure |
| 380 | Seizure like phenomena | Seizure |
| 381 | Tonic clonic movements | Seizure |
| 382 | Tonic convulsion | Seizure |
| 383 | Sepsis | Sepsis |
| 384 | Multiple organ dysfunction syndrome | Shock |
| 385 | Cellulitis | Skin infection |
| 386 | Hypersomnia | Sleep disorder |
| 387 | Insomnia | Sleep disorder |
| 388 | Middle insomnia | Sleep disorder |
| 389 | Poor quality sleep | Sleep disorder |
| 390 | Sleep disorder | Sleep disorder |
| 391 | Anosmia | Smell disorder |
| 392 | Parosmia | Smell disorder |
| 393 | Aphasia | Speech disorder |
| 394 | Aphonia | Speech disorder |
| 395 | Dysarthria | Speech disorder |
| 396 | Dysphonia | Speech disorder |
| 397 | Speech disorder | Speech disorder |
| 398 | Heart rate increased | Tachycardia |

| | | |
|---|---|---|
| 399 | Palpitations | Tachycardia |
| 400 | Tachycardia | Tachycardia |
| 401 | Ageusia | Taste disorder |
| 402 | Dysgeusia | Taste disorder |
| 403 | Taste disorder | Taste disorder |
| 404 | Petechiae | Thrombocytopenia |
| 405 | Autoimmune thyroiditis | Thyroiditis |
| 406 | Glossodynia | Tongue pain |
| 407 | Swollen tongue | Tongue pain |
| 408 | Tongue disorder | Tongue pain |
| 409 | Toothache | Toothache |
| 410 | Face injury | Trauma |
| 411 | Head injury | Trauma |
| 412 | Skin laceration | Trauma |
| 413 | Tooth fracture | Trauma |
| 414 | Pollakiuria | Urinary frequency |
| 415 | Urinary incontinence | Urinary frequency |
| 416 | Bacterial vaginosis | Vaginal infection |
| 417 | Blindness | Visual changes |
| 418 | Blindness transient | Visual changes |
| 419 | Blindness unilateral | Visual changes |
| 420 | Diplopia | Visual changes |
| 421 | Eye movement disorder | Visual changes |
| 422 | Gaze palsy | Visual changes |
| 423 | Mydriasis | Visual changes |
| 424 | Optic neuritis | Visual changes |
| 425 | Photophobia | Visual changes |
| 426 | Photopsia | Visual changes |
| 427 | Tunnel vision | Visual changes |
| 428 | Vision blurred | Visual changes |
| 429 | Visual field defect | Visual changes |
| 430 | Visual impairment | Visual changes |
| 431 | Asthenia | Weakness |
| 432 | Exercise tolerance decreased | Weakness |
| 433 | Grip strength decreased | Weakness |
| 434 | Hypokinesia | Weakness |
| 435 | Hypotonia | Weakness |
| 436 | Mobility decreased | Weakness |
| 437 | Movement disorder | Weakness |
| 438 | Muscular weakness | Weakness |
| 439 | Posture abnormal | Weakness |
| 440 | Weight bearing difficulty | Weakness |
| 441 | Fluid retention | Weight gain |

| State | Pfizer Vaccine | Moderna Vaccine | Janssen Vaccine | Unknown COVID Vaccine | Flu Vaccine |
|---|---|---|---|---|---|
| NA | 9207 (9.1) | NA () | NA () | NA () | NA () |
| AK | 373 (0.4) | 423 (0.3) | 70 (0.3) | 1 (0.2) | 18 (0.4) |
| AL | 878 (0.9) | 1213 (1) | 172 (0.6) | 1 (0.2) | 64 (1.4) |
| AR | 635 (0.6) | 714 (0.6) | 138 (0.5) | 6 (1.3) | 29 (0.6) |
| AZ | 2460 (2.4) | 2540 (2.1) | 512 (1.9) | 12 (2.6) | 116 (2.5) |
| CA | 10725 (10.6) | 12478 (10.3) | 2731 (10.1) | 39 (8.6) | 384 (8.4) |
| CO | 1892 (1.9) | 2533 (2.1) | 563 (2.1) | 6 (1.3) | 119 (2.6) |
| CT | 1237 (1.2) | 1799 (1.5) | 445 (1.7) | 5 (1.1) | 57 (1.3) |
| DC | 286 (0.3) | 279 (0.2) | 99 (0.4) | 1 (0.2) | 5 (0.1) |
| DE | 257 (0.3) | 375 (0.3) | 90 (0.3) | - | - |
| FL | 5751 (5.7) | 7558 (6.2) | 1542 (5.7) | 25 (5.5) | 314 (6.9) |
| GA | 2317 (2.3) | 2988 (2.5) | 577 (2.1) | 10 (2.2) | 138 (3) |
| GU | 12 (0) | 28 (0) | 4 (0) | - | - |
| HI | 523 (0.5) | 450 (0.4) | 95 (0.4) | 1 (0.2) | 17 (0.4) |
| IA | 832 (0.8) | 1094 (0.9) | 254 (0.9) | 3 (0.7) | 43 (0.9) |
| ID | 507 (0.5) | 596 (0.5) | 125 (0.5) | 2 (0.4) | 38 (0.8) |
| IL | 3650 (3.6) | 3839 (3.2) | 1034 (3.8) | 13 (2.9) | 152 (3.3) |
| IN | 3355 (3.3) | 3664 (3) | 1263 (4.7) | 51 (11.2) | 72 (1.6) |
| KS | 855 (0.8) | 1007 (0.8) | 203 (0.8) | 5 (1.1) | 68 (1.5) |
| KY | 1198 (1.2) | 1461 (1.2) | 297 (1.1) | 5 (1.1) | 59 (1.3) |
| LA | 837 (0.8) | 929 (0.8) | 213 (0.8) | 3 (0.7) | 26 (0.6) |
| MA | 2295 (2.3) | 3252 (2.7) | 619 (2.3) | 3 (0.7) | 123 (2.7) |
| MD | 2011 (2) | 2577 (2.1) | 627 (2.3) | 6 (1.3) | 89 (2) |
| ME | 462 (0.5) | 743 (0.6) | 177 (0.7) | 6 (1.3) | 13 (0.3) |
| MH | 3 (0) | 1 (0) | - | - | - |
| MI | 3186 (3.2) | 3550 (2.9) | 772 (2.9) | 16 (3.5) | 141 (3.1) |
| MN | 2094 (2.1) | 2236 (1.8) | 674 (2.5) | 9 (2) | 64 (1.4) |
| MO | 1835 (1.8) | 1967 (1.6) | 445 (1.7) | 4 (0.9) | 108 (2.4) |
| MP | 7 (0) | 9 (0) | 1 (0) | - | - |
| MS | 384 (0.4) | 633 (0.5) | 97 (0.4) | 3 (0.7) | 17 (0.4) |
| MT | 386 (0.4) | 539 (0.4) | 96 (0.4) | 4 (0.9) | 11 (0.2) |
| NC | 2769 (2.7) | 2973 (2.4) | 981 (3.6) | 6 (1.3) | 131 (2.9) |
| ND | 212 (0.2) | 332 (0.3) | 53 (0.2) | - | - |
| NE | 473 (0.5) | 631 (0.5) | 148 (0.5) | 1 (0.2) | 18 (0.4) |
| NH | 486 (0.5) | 734 (0.6) | 173 (0.6) | 2 (0.4) | 21 (0.5) |
| NJ | 2633 (2.6) | 3585 (2.9) | 682 (2.5) | 5 (1.1) | 117 (2.6) |
| NM | 605 (0.6) | 761 (0.6) | 198 (0.7) | 5 (1.1) | 34 (0.7) |
| NV | 643 (0.6) | 960 (0.8) | 224 (0.8) | 4 (0.9) | 42 (0.9) |
| NY | 5955 (5.9) | 6455 (5.3) | 1433 (5.3) | 25 (5.5) | 225 (4.9) |
| OH | 2884 (2.9) | 4470 (3.7) | 933 (3.5) | 12 (2.6) | 212 (4.7) |
| OK | 877 (0.9) | 1153 (0.9) | 195 (0.7) | 2 (0.4) | 30 (0.7) |
| OR | 1308 (1.3) | 1908 (1.6) | 401 (1.5) | 9 (2) | 67 (1.5) |
| PA | 4500 (4.5) | 4541 (3.7) | 965 (3.6) | 11 (2.4) | 227 (5) |
| PR | 432 (0.4) | 453 (0.4) | 72 (0.3) | - | - |
| RI | 351 (0.3) | 466 (0.4) | 122 (0.5) | - | - |
| SC | 1008 (1) | 1200 (1) | 268 (1) | 5 (1.1) | 60 (1.3) |

| | | | | | |
|---|---|---|---|---|---|
| SD | 208 (0.2) | 332 (0.3) | 57 (0.2) | - | - |
| TN | 1551 (1.5) | 1898 (1.6) | 339 (1.3) | 5 (1.1) | 89 (2) |
| TX | 4987 (4.9) | 8257 (6.8) | 1646 (6.1) | 41 (9) | 275 (6) |
| UT | 635 (0.6) | 1004 (0.8) | 233 (0.9) | 7 (1.5) | 47 (1) |
| VA | 2602 (2.6) | 2995 (2.5) | 723 (2.7) | 12 (2.6) | 138 (3) |
| VI | 9 (0) | 13 (0) | - | - | - |
| VT | 227 (0.2) | 394 (0.3) | 118 (0.4) | - | - |
| WA | 2430 (2.4) | 2979 (2.5) | 806 (3) | 12 (2.6) | 157 (3.4) |
| WI | 1960 (1.9) | 2349 (1.9) | 647 (2.4) | 15 (3.3) | 79 (1.7) |
| WV | 408 (0.4) | 559 (0.5) | 76 (0.3) | 2 (0.4) | 28 (0.6) |
| WY | 136 (0.1) | 214 (0.2) | 34 (0.1) | - | - |
| Unknown | 13 (0) | 12 (0) | 1 (0) | - | - |

| 12-15 | Pfizer COVID Vaccine | | Moderna COVID Vaccine | | Janssen Vaccine | Unknown COVID Vaccine | | Flu Vaccine |
|---|---|---|---|---|---|---|---|---|
| **Adverse Events** | **Dose 1 (N = 4,294)** | **Dose 2 (N= 145)** | **Dose 1 (N = 415)** | **Dose 2 (N = 6)** | **Dose 1 (N = 117)** | **Dose 1 (N = 1)** | **Dose 2 (N = 0)** | **Dose 1 (N = 189)** |
| Altered_level_of_consciousness | 506 (11) | 5 (3) | 1 (0) | 0 (0) | 0 (0) | 0 (0) | 0 (0) | 43 (22) |
| Altered_sensation | 1 (0) | 0 (0) | 0 (0) | 0 (0) | 0 (0) | 0 (0) | 0 (0) | 0 (0) |
| Anaemia | 2 (0) | 1 (0) | 0 (0) | 0 (0) | 0 (0) | 0 (0) | 0 (0) | 0 (0) |
| Anaphylaxis | 21 (0) | 0 (0) | 0 (0) | 0 (0) | 0 (0) | 0 (0) | 0 (0) | 0 (0) |
| Anorexia | 23 (0) | 6 (4) | 0 (0) | 0 (0) | 0 (0) | 0 (0) | 0 (0) | 1 (0) |
| Asthma_exacerbation | 8 (0) | 0 (0) | 1 (0) | 0 (0) | 0 (0) | 0 (0) | 0 (0) | 0 (0) |
| Autonomic_neuropathy | 0 (0) | 0 (0) | 0 (0) | 0 (0) | 0 (0) | 0 (0) | 0 (0) | 1 (0) |
| Axillary_pain | 7 (0) | 1 (0) | 0 (0) | 0 (0) | 0 (0) | 0 (0) | 0 (0) | 0 (0) |
| Bacterial_infection_NOS | 1 (0) | 0 (0) | 0 (0) | 0 (0) | 0 (0) | 0 (0) | 0 (0) | 1 (0) |
| Bleeding_NOS | 8 (0) | 0 (0) | 0 (0) | 0 (0) | 0 (0) | 0 (0) | 0 (0) | 1 (0) |
| Bowel_pattern_changes | 43 (1) | 4 (2) | 0 (0) | 0 (0) | 0 (0) | 0 (0) | 0 (0) | 0 (0) |
| Bradycardia | 24 (0) | 0 (0) | 0 (0) | 0 (0) | 0 (0) | 0 (0) | 0 (0) | 2 (1) |
| Cardiorespiratory_arrest | 2 (0) | 0 (0) | 0 (0) | 0 (0) | 0 (0) | 0 (0) | 0 (0) | 0 (0) |
| Central_neuritis | 2 (0) | 0 (0) | 0 (0) | 0 (0) | 0 (0) | 0 (0) | 0 (0) | 1 (0) |
| Central_neuropathy | 1062 (24) | 15 (10) | 5 (1) | 0 (0) | 3 (2) | 0 (0) | 0 (0) | 53 (28) |
| Cerebral_thrombosis_infarction | 2 (0) | 0 (0) | 0 (0) | 0 (0) | 0 (0) | 0 (0) | 0 (0) | 0 (0) |
| Chest_pain | 234 (5) | 45 (31) | 2 (0) | 0 (0) | 0 (0) | 1 (100) | 0 (0) | 0 (0) |
| Choking | 3 (0) | 0 (0) | 0 (0) | 0 (0) | 0 (0) | 0 (0) | 0 (0) | 0 (0) |
| Cognitive_disorder | 75 (1) | 0 (0) | 0 (0) | 0 (0) | 0 (0) | 0 (0) | 0 (0) | 4 (2) |
| Cough | 36 (0) | 1 (0) | 0 (0) | 0 (0) | 0 (0) | 0 (0) | 0 (0) | 1 (0) |
| Death | 2 (0) | 0 (0) | 0 (0) | 0 (0) | 0 (0) | 0 (0) | 0 (0) | 0 (0) |
| Dehydration | 11 (0) | 0 (0) | 1 (0) | 0 (0) | 0 (0) | 0 (0) | 0 (0) | 2 (1) |
| Dermatitis_NOS | 251 (5) | 12 (8) | 1 (0) | 0 (0) | 0 (0) | 0 (0) | 0 (0) | 13 (6) |
| Dizziness | 5 (0) | 0 (0) | 0 (0) | 0 (0) | 0 (0) | 0 (0) | 0 (0) | 5 (2) |
| Drooling | 5 (0) | 1 (0) | 0 (0) | 0 (0) | 0 (0) | 0 (0) | 0 (0) | 0 (0) |
| Dry_mouth | 4 (0) | 0 (0) | 0 (0) | 0 (0) | 0 (0) | 0 (0) | 0 (0) | 0 (0) |
| Dysphagia | 44 (1) | 0 (0) | 0 (0) | 0 (0) | 0 (0) | 0 (0) | 0 (0) | 3 (1) |
| Dyspnea_NOS | 226 (5) | 20 (13) | 2 (0) | 0 (0) | 0 (0) | 0 (0) | 0 (0) | 9 (4) |
| Ear_pain_discomfort | 12 (0) | 1 (0) | 0 (0) | 0 (0) | 0 (0) | 0 (0) | 0 (0) | 0 (0) |
| Edema | 79 (1) | 5 (3) | 2 (0) | 0 (0) | 0 (0) | 0 (0) | 0 (0) | 8 (4) |
| Endocrine_disorder | 7 (0) | 0 (0) | 0 (0) | 0 (0) | 0 (0) | 0 (0) | 0 (0) | 0 (0) |

| | | | | | | | | |
|---|---|---|---|---|---|---|---|---|
| Erythema_NOS | 54 (1) | 1 (0) | 0 (0) | 0 (0) | 0 (0) | 0 (0) | 0 (0) | 7 (3) |
| Eye_irritation | 32 (0) | 2 (1) | 1 (0) | 0 (0) | 1 (0) | 0 (0) | 0 (0) | 1 (0) |
| Facial_neuropathy | 9 (0) | 1 (0) | 0 (0) | 0 (0) | 0 (0) | 0 (0) | 0 (0) | 0 (0) |
| Failure_to_thrive | 80 (1) | 0 (0) | 0 (0) | 0 (0) | 0 (0) | 0 (0) | 0 (0) | 1 (0) |
| Fatigue | 232 (5) | 16 (11) | 0 (0) | 0 (0) | 3 (2) | 0 (0) | 0 (0) | 12 (6) |
| Fever | 645 (15) | 41 (28) | 6 (1) | 0 (0) | 4 (3) | 0 (0) | 0 (0) | 28 (14) |
| Flushing | 125 (2) | 0 (0) | 0 (0) | 0 (0) | 0 (0) | 0 (0) | 0 (0) | 1 (0) |
| Focal_musculoskeletal_pain | 3 (0) | 0 (0) | 0 (0) | 0 (0) | 0 (0) | 0 (0) | 0 (0) | 1 (0) |
| Focal_skin_abnormality | 5 (0) | 0 (0) | 0 (0) | 0 (0) | 0 (0) | 0 (0) | 0 (0) | 0 (0) |
| Groin_pain | 2 (0) | 1 (0) | 0 (0) | 0 (0) | 0 (0) | 0 (0) | 0 (0) | 0 (0) |
| Guillain_Barre_syndrome | 2 (0) | 0 (0) | 0 (0) | 0 (0) | 0 (0) | 0 (0) | 0 (0) | 1 (0) |
| Gynecologic_changes | 25 (0) | 1 (0) | 0 (0) | 0 (0) | 0 (0) | 0 (0) | 0 (0) | 0 (0) |
| Head_trauma | 4 (0) | 0 (0) | 0 (0) | 0 (0) | 0 (0) | 0 (0) | 0 (0) | 1 (0) |
| Headache | 332 (7) | 27 (18) | 3 (0) | 1 (16) | 5 (4) | 0 (0) | 0 (0) | 15 (7) |
| Hearing_changes | 67 (1) | 0 (0) | 1 (0) | 0 (0) | 0 (0) | 0 (0) | 0 (0) | 3 (1) |
| Hematuria | 2 (0) | 0 (0) | 0 (0) | 0 (0) | 0 (0) | 0 (0) | 0 (0) | 1 (0) |
| Hepatitis | 3 (0) | 1 (0) | 0 (0) | 0 (0) | 0 (0) | 0 (0) | 0 (0) | 1 (0) |
| Hypertension | 6 (0) | 0 (0) | 0 (0) | 0 (0) | 0 (0) | 0 (0) | 0 (0) | 0 (0) |
| Hypotension | 778 (18) | 5 (3) | 5 (1) | 0 (0) | 1 (0) | 0 (0) | 0 (0) | 62 (32) |
| Hypothermia | 5 (0) | 0 (0) | 0 (0) | 0 (0) | 0 (0) | 0 (0) | 0 (0) | 2 (1) |
| Hypovolemia | 7 (0) | 0 (0) | 0 (0) | 0 (0) | 0 (0) | 0 (0) | 0 (0) | 0 (0) |
| Injection_site_complication | 185 (4) | 9 (6) | 8 (1) | 0 (0) | 1 (0) | 0 (0) | 0 (0) | 12 (6) |
| Irregular_heart_rate | 8 (0) | 1 (0) | 0 (0) | 0 (0) | 0 (0) | 0 (0) | 0 (0) | 0 (0) |
| Lymphadenopathy | 66 (1) | 6 (4) | 0 (0) | 0 (0) | 0 (0) | 0 (0) | 0 (0) | 2 (1) |
| Malaise | 165 (3) | 7 (4) | 0 (0) | 0 (0) | 0 (0) | 0 (0) | 0 (0) | 11 (5) |
| Memory_disorder | 9 (0) | 0 (0) | 0 (0) | 0 (0) | 0 (0) | 0 (0) | 0 (0) | 1 (0) |
| Myocarditis | 41 (0) | 16 (11) | 0 (0) | 0 (0) | 0 (0) | 0 (0) | 0 (0) | 0 (0) |
| Nausea_vomiting | 583 (13) | 23 (15) | 2 (0) | 0 (0) | 0 (0) | 0 (0) | 0 (0) | 26 (13) |
| Neck_back_pain | 30 (0) | 6 (4) | 0 (0) | 0 (0) | 1 (0) | 0 (0) | 0 (0) | 3 (1) |
| Neuropathy_NOS | 10 (0) | 1 (0) | 0 (0) | 0 (0) | 0 (0) | 0 (0) | 0 (0) | 0 (0) |
| Nonspecific_musculoskeletal_pain | 214 (4) | 26 (17) | 6 (1) | 0 (0) | 1 (0) | 0 (0) | 0 (0) | 10 (5) |
| Nosebleed | 25 (0) | 0 (0) | 1 (0) | 1 (16) | 0 (0) | 0 (0) | 0 (0) | 1 (0) |
| Other_viral_infection | 10 (0) | 1 (0) | 0 (0) | 0 (0) | 0 (0) | 0 (0) | 0 (0) | 0 (0) |
| Overdose | 6 (0) | 0 (0) | 0 (0) | 0 (0) | 0 (0) | 0 (0) | 0 (0) | 0 (0) |
| Pallor | 351 (8) | 6 (4) | 2 (0) | 0 (0) | 0 (0) | 0 (0) | 0 (0) | 24 (12) |

| | | | | | | | | |
|---|---|---|---|---|---|---|---|---|
| Pericarditis | 14 (0) | 4 (2) | 0 (0) | 0 (0) | 0 (0) | 0 (0) | 0 (0) | 0 (0) |
| Perioral_inflammation | 43 (1) | 0 (0) | 0 (0) | 0 (0) | 0 (0) | 0 (0) | 0 (0) | 2 (1) |
| Peripheral_neuropathy | 122 (2) | 2 (1) | 0 (0) | 0 (0) | 0 (0) | 0 (0) | 0 (0) | 10 (5) |
| Pharyngitis | 112 (2) | 2 (1) | 1 (0) | 0 (0) | 0 (0) | 0 (0) | 0 (0) | 6 (3) |
| Pneumonia | 6 (0) | 1 (0) | 0 (0) | 0 (0) | 0 (0) | 0 (0) | 0 (0) | 0 (0) |
| Psychiatric_amplification | 127 (2) | 2 (1) | 0 (0) | 0 (0) | 1 (0) | 0 (0) | 0 (0) | 7 (3) |
| Psychiatric_depression | 14 (0) | 1 (0) | 0 (0) | 0 (0) | 0 (0) | 0 (0) | 0 (0) | 0 (0) |
| Psychiatric_disorder | 1 (0) | 0 (0) | 0 (0) | 0 (0) | 0 (0) | 0 (0) | 0 (0) | 0 (0) |
| Pulmonary_embolism | 1 (0) | 1 (0) | 0 (0) | 0 (0) | 0 (0) | 0 (0) | 0 (0) | 0 (0) |
| Rash | 119 (2) | 5 (3) | 0 (0) | 0 (0) | 0 (0) | 0 (0) | 0 (0) | 10 (5) |
| Raynauds_phenomenon | 0 (0) | 0 (0) | 0 (0) | 0 (0) | 0 (0) | 0 (0) | 0 (0) | 1 (0) |
| Renal_failure | 3 (0) | 0 (0) | 0 (0) | 0 (0) | 0 (0) | 0 (0) | 0 (0) | 0 (0) |
| Respiratory_failure | 10 (0) | 1 (0) | 0 (0) | 0 (0) | 0 (0) | 0 (0) | 0 (0) | 0 (0) |
| Rhinitis | 31 (0) | 1 (0) | 0 (0) | 0 (0) | 0 (0) | 0 (0) | 0 (0) | 0 (0) |
| Seizure | 130 (3) | 6 (4) | 0 (0) | 0 (0) | 1 (0) | 0 (0) | 0 (0) | 14 (7) |
| Sepsis | 1 (0) | 0 (0) | 0 (0) | 0 (0) | 0 (0) | 0 (0) | 0 (0) | 0 (0) |
| Skin_infection | 1 (0) | 0 (0) | 0 (0) | 0 (0) | 0 (0) | 0 (0) | 0 (0) | 2 (1) |
| Smell_disorder | 36 (0) | 6 (4) | 0 (0) | 0 (0) | 0 (0) | 0 (0) | 0 (0) | 4 (2) |
| Speech_disorder | 34 (0) | 1 (0) | 0 (0) | 0 (0) | 0 (0) | 0 (0) | 0 (0) | 2 (1) |
| Tachycardia | 99 (2) | 8 (5) | 0 (0) | 0 (0) | 0 (0) | 0 (0) | 0 (0) | 2 (1) |
| Taste_disorder | 10 (0) | 1 (0) | 0 (0) | 0 (0) | 0 (0) | 0 (0) | 0 (0) | 0 (0) |
| Thrombocytopenia | 15 (0) | 2 (1) | 0 (0) | 0 (0) | 0 (0) | 0 (0) | 0 (0) | 0 (0) |
| Thyroiditis | 1 (0) | 0 (0) | 0 (0) | 0 (0) | 0 (0) | 0 (0) | 0 (0) | 1 (0) |
| Tongue_pain | 17 (0) | 0 (0) | 0 (0) | 0 (0) | 1 (0) | 0 (0) | 0 (0) | 1 (0) |
| Toothache | 1 (0) | 0 (0) | 0 (0) | 0 (0) | 0 (0) | 0 (0) | 0 (0) | 0 (0) |
| Trauma | 81 (1) | 1 (0) | 1 (0) | 0 (0) | 0 (0) | 0 (0) | 0 (0) | 8 (4) |
| Urinary_frequency | 5 (0) | 0 (0) | 0 (0) | 0 (0) | 0 (0) | 0 (0) | 0 (0) | 1 (0) |
| Visual_changes | 243 (5) | 2 (1) | 2 (0) | 0 (0) | 0 (0) | 0 (0) | 0 (0) | 22 (11) |
| Weakness | 168 (3) | 6 (4) | 1 (0) | 1 (16) | 1 (0) | 0 (0) | 0 (0) | 14 (7) |
| **16-30** | | | | | | | | |
| **Adverse Events** | **Dose 1 (N = 17,875)** | **Dose 2 (N = 947)** | **Dose 1 (N = 18,910)** | **Dose 2 (N = 769)** | **Dose 1 (N = 6,761)** | **Dose 1 (N = 87)** | **Dose 2 (N = 0)** | **Dose 1 (N = 607)** |
| Alopecia | 13 (0) | 2 (0) | 9 (0) | 2 (0) | 8 (0) | 0 (0) | 0 (0) | 2 (0) |
| Altered_level_of_consciousness | 1669 (9) | 14 (1) | 1006 (5) | 18 (2) | 802 (11) | 3 (3) | 0 (0) | 87 (14) |
| Altered_sensation | 7 (0) | 0 (0) | 17 (0) | 0 (0) | 17 (0) | 0 (0) | 0 (0) | 1 (0) |

| | | | | | | | | |
|---|---|---|---|---|---|---|---|---|
| Anaemia | 10 (0) | 2 (0) | 3 (0) | 1 (0) | 6 (0) | 0 (0) | 0 (0) | 0 (0) |
| Anaphylaxis | 105 (0) | 4 (0) | 69 (0) | 0 (0) | 11 (0) | 1 (1) | 0 (0) | 5 (0) |
| Anorexia | 160 (0) | 17 (1) | 183 (0) | 10 (1) | 120 (1) | 0 (0) | 0 (0) | 2 (0) |
| Arthropathy | 4 (0) | 0 (0) | 2 (0) | 0 (0) | 1 (0) | 0 (0) | 0 (0) | 1 (0) |
| Asthma_exacerbation | 24 (0) | 2 (0) | 26 (0) | 1 (0) | 9 (0) | 1 (1) | 0 (0) | 0 (0) |
| Autonomic_neuropathy | 2 (0) | 0 (0) | 2 (0) | 0 (0) | 1 (0) | 0 (0) | 0 (0) | 0 (0) |
| Axillary_pain | 98 (0) | 8 (0) | 205 (1) | 6 (0) | 3 (0) | 0 (0) | 0 (0) | 1 (0) |
| Bacterial_infection_NOS | 27 (0) | 3 (0) | 14 (0) | 1 (0) | 4 (0) | 0 (0) | 0 (0) | 1 (0) |
| Bleeding_NOS | 35 (0) | 4 (0) | 18 (0) | 0 (0) | 10 (0) | 0 (0) | 0 (0) | 3 (0) |
| Bowel_pattern_changes | 421 (2) | 39 (4) | 366 (1) | 28 (3) | 180 (2) | 1 (1) | 0 (0) | 9 (1) |
| Bradycardia | 93 (0) | 5 (0) | 66 (0) | 0 (0) | 50 (0) | 1 (1) | 0 (0) | 4 (0) |
| Breast_pain | 22 (0) | 4 (0) | 13 (0) | 1 (0) | 4 (0) | 0 (0) | 0 (0) | 0 (0) |
| COPD_exacerbation | 1 (0) | 0 (0) | 0 (0) | 0 (0) | 0 (0) | 0 (0) | 0 (0) | 0 (0) |
| Cardiorespiratory_arrest | 13 (0) | 0 (0) | 3 (0) | 0 (0) | 1 (0) | 0 (0) | 0 (0) | 0 (0) |
| Central_neuritis | 2 (0) | 0 (0) | 3 (0) | 0 (0) | 1 (0) | 0 (0) | 0 (0) | 0 (0) |
| Central_neuropathy | 4799 (26) | 108 (11) | 3152 (16) | 76 (9) | 2109 (31) | 14 (16) | 0 (0) | 145 (23) |
| Cerebral_hemorrhage | 4 (0) | 1 (0) | 3 (0) | 0 (0) | 1 (0) | 0 (0) | 0 (0) | 0 (0) |
| Cerebral_thrombosis_infarction | 21 (0) | 2 (0) | 12 (0) | 0 (0) | 9 (0) | 0 (0) | 0 (0) | 1 (0) |
| Chest_pain | 1106 (6) | 114 (12) | 698 (3) | 67 (8) | 312 (4) | 4 (4) | 0 (0) | 7 (1) |
| Choking | 6 (0) | 0 (0) | 6 (0) | 1 (0) | 1 (0) | 0 (0) | 0 (0) | 2 (0) |
| Cognitive_disorder | 231 (1) | 6 (0) | 124 (0) | 3 (0) | 106 (1) | 0 (0) | 0 (0) | 5 (0) |
| Congestive_heart_failure | 1 (0) | 1 (0) | 0 (0) | 0 (0) | 0 (0) | 0 (0) | 0 (0) | 0 (0) |
| Cough | 404 (2) | 38 (4) | 276 (1) | 28 (3) | 98 (1) | 2 (2) | 0 (0) | 7 (1) |
| Death | 14 (0) | 1 (0) | 18 (0) | 2 (0) | 3 (0) | 0 (0) | 0 (0) | 0 (0) |
| Deep_vein_thrombosis | 20 (0) | 7 (0) | 11 (0) | 3 (0) | 10 (0) | 0 (0) | 0 (0) | 0 (0) |
| Dehydration | 71 (0) | 2 (0) | 44 (0) | 3 (0) | 51 (0) | 1 (1) | 0 (0) | 0 (0) |
| Dermatitis_NOS | 1695 (9) | 95 (10) | 2372 (12) | 67 (8) | 310 (4) | 3 (3) | 0 (0) | 49 (8) |
| Dizziness | 63 (0) | 6 (0) | 40 (0) | 4 (0) | 32 (0) | 0 (0) | 0 (0) | 2 (0) |
| Drooling | 5 (0) | 0 (0) | 2 (0) | 0 (0) | 3 (0) | 0 (0) | 0 (0) | 0 (0) |
| Dry_mouth | 64 (0) | 1 (0) | 37 (0) | 3 (0) | 17 (0) | 0 (0) | 0 (0) | 2 (0) |
| Dysphagia | 202 (1) | 6 (0) | 149 (0) | 4 (0) | 33 (0) | 0 (0) | 0 (0) | 3 (0) |
| Dyspnea_NOS | 1414 (7) | 101 (10) | 984 (5) | 69 (8) | 464 (6) | 10 (11) | 0 (0) | 23 (3) |
| Ear_pain_discomfort | 147 (0) | 13 (1) | 86 (0) | 8 (1) | 27 (0) | 1 (1) | 0 (0) | 1 (0) |
| Edema | 583 (3) | 35 (3) | 1011 (5) | 31 (4) | 152 (2) | 1 (1) | 0 (0) | 36 (5) |
| Endocrine_disorder | 123 (0) | 7 (0) | 108 (0) | 8 (1) | 70 (1) | 1 (1) | 0 (0) | 1 (0) |

| | | | | | | | | |
|---|---|---|---|---|---|---|---|---|
| Erythema_NOS | 361 (2) | 14 (1) | 808 (4) | 22 (2) | 92 (1) | 2 (2) | 0 (0) | 36 (5) |
| Eye_irritation | 227 (1) | 12 (1) | 185 (0) | 13 (1) | 88 (1) | 0 (0) | 0 (0) | 10 (1) |
| Facial_neuropathy | 143 (0) | 13 (1) | 109 (0) | 12 (1) | 36 (0) | 0 (0) | 0 (0) | 2 (0) |
| Failure_to_thrive | 366 (2) | 46 (4) | 354 (1) | 21 (2) | 157 (2) | 0 (0) | 0 (0) | 14 (2) |
| Fatigue | 2161 (12) | 181 (19) | 2115 (11) | 125 (16) | 1130 (16) | 11 (12) | 0 (0) | 22 (3) |
| Fetal_death | 49 (0) | 14 (1) | 41 (0) | 11 (1) | 10 (0) | 0 (0) | 0 (0) | 2 (0) |
| Fever | 4130 (23) | 242 (25) | 4158 (21) | 197 (25) | 3070 (45) | 32 (36) | 0 (0) | 102 (16) |
| Flushing | 617 (3) | 7 (0) | 349 (1) | 2 (0) | 145 (2) | 0 (0) | 0 (0) | 7 (1) |
| Focal_musculoskeletal_pain | 42 (0) | 2 (0) | 33 (0) | 1 (0) | 15 (0) | 0 (0) | 0 (0) | 6 (0) |
| Focal_skin_abnormality | 39 (0) | 1 (0) | 36 (0) | 1 (0) | 3 (0) | 0 (0) | 0 (0) | 0 (0) |
| Gastrointestinal_bleeding | 1 (0) | 0 (0) | 2 (0) | 0 (0) | 2 (0) | 0 (0) | 0 (0) | 0 (0) |
| Generalized_allergic_response | 3 (0) | 0 (0) | 1 (0) | 0 (0) | 0 (0) | 0 (0) | 0 (0) | 0 (0) |
| Groin_pain | 6 (0) | 0 (0) | 7 (0) | 0 (0) | 4 (0) | 0 (0) | 0 (0) | 0 (0) |
| Guillain_Barre_syndrome | 6 (0) | 1 (0) | 7 (0) | 4 (0) | 5 (0) | 0 (0) | 0 (0) | 3 (0) |
| Gynecologic_changes | 357 (1) | 67 (7) | 211 (1) | 43 (5) | 89 (1) | 0 (0) | 0 (0) | 0 (0) |
| Head_trauma | 11 (0) | 0 (0) | 9 (0) | 0 (0) | 5 (0) | 0 (0) | 0 (0) | 0 (0) |
| Headache | 2599 (14) | 194 (20) | 2583 (13) | 166 (21) | 1958 (28) | 20 (22) | 0 (0) | 38 (6) |
| Hearing_changes | 411 (2) | 28 (2) | 208 (1) | 13 (1) | 153 (2) | 3 (3) | 0 (0) | 11 (1) |
| Hematuria | 23 (0) | 4 (0) | 13 (0) | 2 (0) | 7 (0) | 0 (0) | 0 (0) | 0 (0) |
| Hepatitis | 15 (0) | 6 (0) | 14 (0) | 2 (0) | 3 (0) | 0 (0) | 0 (0) | 0 (0) |
| Hypertension | 96 (0) | 4 (0) | 69 (0) | 6 (0) | 26 (0) | 0 (0) | 0 (0) | 2 (0) |
| Hypotension | 2320 (12) | 20 (2) | 1314 (6) | 15 (1) | 969 (14) | 9 (10) | 0 (0) | 102 (16) |
| Hypothermia | 10 (0) | 0 (0) | 10 (0) | 1 (0) | 6 (0) | 0 (0) | 0 (0) | 1 (0) |
| Hypovolemia | 35 (0) | 1 (0) | 31 (0) | 0 (0) | 24 (0) | 0 (0) | 0 (0) | 0 (0) |
| Injection_site_complication | 1687 (9) | 61 (6) | 4259 (22) | 81 (10) | 542 (8) | 4 (4) | 0 (0) | 121 (19) |
| Irregular_heart_rate | 56 (0) | 7 (0) | 34 (0) | 2 (0) | 6 (0) | 0 (0) | 0 (0) | 0 (0) |
| Lymphadenopathy | 507 (2) | 45 (4) | 659 (3) | 27 (3) | 38 (0) | 0 (0) | 0 (0) | 6 (0) |
| Malaise | 995 (5) | 66 (6) | 705 (3) | 42 (5) | 422 (6) | 5 (5) | 0 (0) | 26 (4) |
| Memory_disorder | 60 (0) | 6 (0) | 34 (0) | 3 (0) | 22 (0) | 0 (0) | 0 (0) | 3 (0) |
| Myocardial_infarction | 7 (0) | 2 (0) | 4 (0) | 0 (0) | 0 (0) | 1 (1) | 0 (0) | 0 (0) |
| Myocarditis | 84 (0) | 37 (3) | 51 (0) | 11 (1) | 6 (0) | 0 (0) | 0 (0) | 1 (0) |
| Nausea_vomiting | 2747 (15) | 115 (12) | 2444 (12) | 96 (12) | 1586 (23) | 15 (17) | 0 (0) | 70 (11) |
| Neck_back_pain | 366 (2) | 33 (3) | 329 (1) | 26 (3) | 254 (3) | 3 (3) | 0 (0) | 10 (1) |
| Neuropathy_NOS | 127 (0) | 8 (0) | 135 (0) | 8 (1) | 32 (0) | 0 (0) | 0 (0) | 2 (0) |
| Nonspecific_musculoskeletal_pain | 2245 (12) | 181 (19) | 2834 (14) | 140 (18) | 1143 (16) | 13 (14) | 0 (0) | 96 (15) |

| | | | | | | | | |
|---|---|---|---|---|---|---|---|---|
| Nosebleed | 65 (0) | 6 (0) | 40 (0) | 1 (0) | 29 (0) | 0 (0) | 0 (0) | 2 (0) |
| Other_viral_infection | 82 (0) | 10 (1) | 61 (0) | 7 (0) | 20 (0) | 0 (0) | 0 (0) | 0 (0) |
| Overdose | 5 (0) | 0 (0) | 2 (0) | 0 (0) | 0 (0) | 0 (0) | 0 (0) | 1 (0) |
| Pallor | 800 (4) | 2 (0) | 478 (2) | 2 (0) | 339 (5) | 2 (2) | 0 (0) | 33 (5) |
| Pericarditis | 46 (0) | 18 (1) | 30 (0) | 11 (1) | 10 (0) | 1 (1) | 0 (0) | 0 (0) |
| Perioral_inflammation | 509 (2) | 13 (1) | 362 (1) | 13 (1) | 64 (0) | 1 (1) | 0 (0) | 12 (1) |
| Peripheral_neuropathy | 1530 (8) | 76 (8) | 1174 (6) | 57 (7) | 518 (7) | 4 (4) | 0 (0) | 51 (8) |
| Peripheral_thrombosis | 0 (0) | 0 (0) | 0 (0) | 0 (0) | 1 (0) | 0 (0) | 0 (0) | 0 (0) |
| Pharyngitis | 1014 (5) | 49 (5) | 727 (3) | 27 (3) | 177 (2) | 1 (1) | 0 (0) | 21 (3) |
| Pneumonia | 18 (0) | 2 (0) | 7 (0) | 1 (0) | 6 (0) | 0 (0) | 0 (0) | 1 (0) |
| Pregnancy_complication | 13 (0) | 4 (0) | 12 (0) | 2 (0) | 1 (0) | 0 (0) | 0 (0) | 1 (0) |
| Psychiatric_amplification | 672 (3) | 18 (1) | 464 (2) | 16 (2) | 230 (3) | 1 (1) | 0 (0) | 13 (2) |
| Psychiatric_depression | 63 (0) | 4 (0) | 35 (0) | 0 (0) | 33 (0) | 0 (0) | 0 (0) | 2 (0) |
| Psychiatric_disorder | 1 (0) | 0 (0) | 0 (0) | 0 (0) | 0 (0) | 0 (0) | 0 (0) | 0 (0) |
| Pulmonary_embolism | 28 (0) | 10 (1) | 17 (0) | 6 (0) | 15 (0) | 0 (0) | 0 (0) | 0 (0) |
| Rash | 714 (3) | 30 (3) | 747 (3) | 31 (4) | 130 (1) | 1 (1) | 0 (0) | 41 (6) |
| Raynauds_phenomenon | 2 (0) | 1 (0) | 2 (0) | 0 (0) | 0 (0) | 0 (0) | 0 (0) | 0 (0) |
| Renal_failure | 9 (0) | 2 (0) | 3 (0) | 0 (0) | 0 (0) | 0 (0) | 0 (0) | 0 (0) |
| Respiratory_failure | 25 (0) | 0 (0) | 13 (0) | 0 (0) | 7 (0) | 1 (1) | 0 (0) | 1 (0) |
| Rhinitis | 345 (1) | 30 (3) | 195 (1) | 21 (2) | 76 (1) | 0 (0) | 0 (0) | 3 (0) |
| Seizure | 484 (2) | 10 (1) | 320 (1) | 4 (0) | 200 (2) | 2 (2) | 0 (0) | 26 (4) |
| Sepsis | 4 (0) | 1 (0) | 1 (0) | 0 (0) | 1 (0) | 0 (0) | 0 (0) | 0 (0) |
| Shock | 0 (0) | 0 (0) | 0 (0) | 0 (0) | 1 (0) | 0 (0) | 0 (0) | 0 (0) |
| Skin_infection | 5 (0) | 0 (0) | 70 (0) | 4 (0) | 5 (0) | 0 (0) | 0 (0) | 5 (0) |
| Smell_disorder | 429 (2) | 57 (6) | 366 (1) | 39 (5) | 262 (3) | 2 (2) | 0 (0) | 8 (1) |
| Speech_disorder | 146 (0) | 7 (0) | 97 (0) | 5 (0) | 42 (0) | 0 (0) | 0 (0) | 7 (1) |
| Tachycardia | 1014 (5) | 56 (5) | 703 (3) | 41 (5) | 280 (4) | 7 (8) | 0 (0) | 14 (2) |
| Taste_disorder | 250 (1) | 34 (3) | 147 (0) | 14 (1) | 43 (0) | 0 (0) | 0 (0) | 1 (0) |
| Thrombocytopenia | 29 (0) | 2 (0) | 16 (0) | 3 (0) | 12 (0) | 0 (0) | 0 (0) | 0 (0) |
| Thyroiditis | 0 (0) | 0 (0) | 1 (0) | 0 (0) | 0 (0) | 0 (0) | 0 (0) | 0 (0) |
| Tongue_pain | 190 (1) | 1 (0) | 130 (0) | 3 (0) | 17 (0) | 0 (0) | 0 (0) | 2 (0) |
| Toothache | 5 (0) | 0 (0) | 12 (0) | 0 (0) | 8 (0) | 0 (0) | 0 (0) | 1 (0) |
| Trauma | 249 (1) | 0 (0) | 140 (0) | 0 (0) | 132 (1) | 0 (0) | 0 (0) | 16 (2) |
| Urinary_frequency | 40 (0) | 1 (0) | 27 (0) | 2 (0) | 28 (0) | 0 (0) | 0 (0) | 1 (0) |
| Vaginal_infection | 1 (0) | 0 (0) | 0 (0) | 0 (0) | 1 (0) | 0 (0) | 0 (0) | 0 (0) |

| | | | | | | | | |
|---|---|---|---|---|---|---|---|---|
| Visual_changes | 902 (5) | 24 (2) | 521 (2) | 23 (2) | 431 (6) | 2 (2) | 0 (0) | 35 (5) |
| Weakness | 933 (5) | 64 (6) | 817 (4) | 41 (5) | 504 (7) | 3 (3) | 0 (0) | 52 (8) |
| Weight_gain | 2 (0) | 0 (0) | 2 (0) | 0 (0) | 0 (0) | 0 (0) | 0 (0) | 0 (0) |
| **31-64** | | | | | | | | |
| **Adverse Events** | **Dose 1 (N = 59,688)** | **Dose 2 (N = 4,356)** | **Dose 1 (N = 68,185)** | **Dose 2 (N = 3,186)** | **Dose 1 (N = 17,364)** | **Dose 1 (N = 285)** | **Dose 2 (N = 3)** | **Dose 1 (N = 1,897)** |
| NA | 25338 (42) | 2780 (63) | 21486 (31) | 1891 (59) | 7135 (41) | 114 (40) | 2 (66) | 516 (27) |
| Alopecia | 79 (0) | 18 (0) | 72 (0) | 19 (0) | 30 (0) | 2 (0) | 0 (0) | 1 (0) |
| Altered_level_of_consciousness | 1767 (2) | 87 (1) | 1544 (2) | 63 (1) | 1045 (6) | 12 (4) | 0 (0) | 41 (2) |
| Altered_sensation | 65 (0) | 6 (0) | 72 (0) | 4 (0) | 39 (0) | 1 (0) | 0 (0) | 1 (0) |
| Anaemia | 37 (0) | 11 (0) | 36 (0) | 11 (0) | 15 (0) | 0 (0) | 0 (0) | 1 (0) |
| Anaphylaxis | 374 (0) | 6 (0) | 276 (0) | 5 (0) | 38 (0) | 1 (0) | 0 (0) | 7 (0) |
| Anorexia | 773 (1) | 66 (1) | 955 (1) | 83 (2) | 409 (2) | 7 (2) | 0 (0) | 22 (1) |
| Arthropathy | 21 (0) | 5 (0) | 9 (0) | 2 (0) | 5 (0) | 0 (0) | 0 (0) | 0 (0) |
| Asthma_exacerbation | 180 (0) | 12 (0) | 172 (0) | 9 (0) | 39 (0) | 0 (0) | 0 (0) | 0 (0) |
| Autonomic_neuropathy | 12 (0) | 3 (0) | 8 (0) | 0 (0) | 2 (0) | 1 (0) | 0 (0) | 0 (0) |
| Axillary_pain | 440 (0) | 56 (1) | 880 (1) | 25 (0) | 72 (0) | 1 (0) | 0 (0) | 21 (1) |
| Bacterial_infection_NOS | 83 (0) | 15 (0) | 95 (0) | 8 (0) | 35 (0) | 0 (0) | 0 (0) | 2 (0) |
| Bleeding_NOS | 105 (0) | 15 (0) | 89 (0) | 8 (0) | 27 (0) | 0 (0) | 0 (0) | 8 (0) |
| Bowel_pattern_changes | 2433 (4) | 169 (3) | 2396 (3) | 137 (4) | 715 (4) | 15 (5) | 1 (33) | 41 (2) |
| Bradycardia | 170 (0) | 9 (0) | 146 (0) | 7 (0) | 66 (0) | 0 (0) | 0 (0) | 2 (0) |
| Breast_pain | 201 (0) | 18 (0) | 178 (0) | 21 (0) | 41 (0) | 0 (0) | 0 (0) | 3 (0) |
| COPD_exacerbation | 14 (0) | 5 (0) | 12 (0) | 0 (0) | 4 (0) | 0 (0) | 0 (0) | 0 (0) |
| Cardiorespiratory_arrest | 68 (0) | 12 (0) | 74 (0) | 10 (0) | 21 (0) | 2 (0) | 0 (0) | 0 (0) |
| Central_neuritis | 6 (0) | 3 (0) | 7 (0) | 0 (0) | 3 (0) | 0 (0) | 0 (0) | 0 (0) |
| Central_neuropathy | 11144 (18) | 541 (12) | 9523 (13) | 418 (13) | 4007 (23) | 54 (18) | 0 (0) | 185 (9) |
| Cerebral_hemorrhage | 28 (0) | 3 (0) | 22 (0) | 2 (0) | 31 (0) | 0 (0) | 0 (0) | 0 (0) |
| Cerebral_thrombosis_infarction | 252 (0) | 43 (0) | 223 (0) | 34 (1) | 148 (0) | 4 (1) | 0 (0) | 1 (0) |
| Chest_pain | 3606 (6) | 278 (6) | 2759 (4) | 168 (5) | 923 (5) | 8 (2) | 0 (0) | 38 (2) |
| Choking | 39 (0) | 2 (0) | 33 (0) | 0 (0) | 5 (0) | 0 (0) | 0 (0) | 0 (0) |
| Cognitive_disorder | 539 (0) | 44 (1) | 499 (0) | 32 (1) | 217 (1) | 2 (0) | 0 (0) | 5 (0) |
| Congestive_heart_failure | 22 (0) | 4 (0) | 15 (0) | 4 (0) | 9 (0) | 0 (0) | 0 (0) | 0 (0) |
| Cough | 2162 (3) | 201 (4) | 1628 (2) | 111 (3) | 482 (2) | 7 (2) | 2 (66) | 43 (2) |
| Death | 207 (0) | 40 (0) | 285 (0) | 33 (1) | 102 (0) | 2 (0) | 0 (0) | 5 (0) |
| Deep_vein_thrombosis | 152 (0) | 42 (0) | 156 (0) | 41 (1) | 153 (0) | 4 (1) | 0 (0) | 0 (0) |

| | | | | | | | | |
|---|---|---|---|---|---|---|---|---|
| Dehydration | 189 (0) | 14 (0) | 200 (0) | 14 (0) | 108 (0) | 2 (0) | 0 (0) | 5 (0) |
| Dermatitis_NOS | 7184 (12) | 390 (8) | 12163 (17) | 392 (12) | 1434 (8) | 27 (9) | 0 (0) | 223 (11) |
| Dizziness | 357 (0) | 45 (1) | 266 (0) | 31 (0) | 120 (0) | 3 (1) | 0 (0) | 7 (0) |
| Drooling | 23 (0) | 2 (0) | 19 (0) | 2 (0) | 6 (0) | 0 (0) | 0 (0) | 0 (0) |
| Dry_mouth | 338 (0) | 12 (0) | 300 (0) | 9 (0) | 76 (0) | 1 (0) | 0 (0) | 2 (0) |
| Dysphagia | 893 (1) | 31 (0) | 662 (0) | 17 (0) | 101 (0) | 3 (1) | 0 (0) | 8 (0) |
| Dyspnea_NOS | 4930 (8) | 396 (9) | 4142 (6) | 277 (8) | 1413 (8) | 16 (5) | 2 (66) | 77 (4) |
| Ear_pain_discomfort | 813 (1) | 80 (1) | 580 (0) | 47 (1) | 204 (1) | 3 (1) | 0 (0) | 6 (0) |
| Edema | 3192 (5) | 233 (5) | 5448 (7) | 217 (6) | 777 (4) | 17 (5) | 0 (0) | 206 (10) |
| Endocrine_disorder | 521 (0) | 26 (0) | 398 (0) | 25 (0) | 145 (0) | 1 (0) | 0 (0) | 3 (0) |
| Erythema_NOS | 1767 (2) | 97 (2) | 4487 (6) | 95 (2) | 326 (1) | 8 (2) | 0 (0) | 154 (8) |
| Eye_irritation | 1337 (2) | 98 (2) | 1108 (1) | 64 (2) | 331 (1) | 5 (1) | 0 (0) | 37 (1) |
| Facial_neuropathy | 866 (1) | 79 (1) | 719 (1) | 54 (1) | 201 (1) | 2 (0) | 0 (0) | 15 (0) |
| Failure_to_thrive | 1680 (2) | 200 (4) | 1854 (2) | 153 (4) | 685 (3) | 11 (3) | 0 (0) | 65 (3) |
| Fatigue | 9265 (15) | 743 (17) | 10519 (15) | 659 (20) | 3690 (21) | 66 (23) | 1 (33) | 118 (6) |
| Fetal_death | 146 (0) | 36 (0) | 112 (0) | 27 (0) | 18 (0) | 0 (0) | 0 (0) | 1 (0) |
| Fever | 12655 (21) | 887 (20) | 16178 (23) | 899 (28) | 6855 (39) | 99 (34) | 3 (100) | 335 (17) |
| Flushing | 1689 (2) | 29 (0) | 1312 (1) | 13 (0) | 264 (1) | 0 (0) | 0 (0) | 9 (0) |
| Focal_musculoskeletal_pain | 321 (0) | 39 (0) | 283 (0) | 21 (0) | 94 (0) | 2 (0) | 0 (0) | 28 (1) |
| Focal_skin_abnormality | 236 (0) | 30 (0) | 253 (0) | 17 (0) | 46 (0) | 1 (0) | 0 (0) | 10 (0) |
| Gastrointestinal_bleeding | 19 (0) | 10 (0) | 6 (0) | 1 (0) | 2 (0) | 0 (0) | 0 (0) | 0 (0) |
| Generalized_allergic_response | 8 (0) | 0 (0) | 2 (0) | 0 (0) | 0 (0) | 0 (0) | 0 (0) | 0 (0) |
| Groin_pain | 46 (0) | 5 (0) | 30 (0) | 5 (0) | 24 (0) | 0 (0) | 0 (0) | 2 (0) |
| Guillain_Barre_syndrome | 45 (0) | 9 (0) | 51 (0) | 9 (0) | 49 (0) | 3 (1) | 0 (0) | 10 (0) |
| Gynecologic_changes | 996 (1) | 205 (4) | 630 (0) | 142 (4) | 284 (1) | 3 (1) | 0 (0) | 2 (0) |
| Head_trauma | 8 (0) | 2 (0) | 13 (0) | 0 (0) | 6 (0) | 0 (0) | 0 (0) | 0 (0) |
| Headache | 11623 (19) | 840 (19) | 13229 (19) | 724 (22) | 5580 (32) | 102 (35) | 0 (0) | 176 (9) |
| Hearing_changes | 2054 (3) | 245 (5) | 1471 (2) | 181 (5) | 486 (2) | 9 (3) | 0 (0) | 15 (0) |
| Hematuria | 67 (0) | 11 (0) | 75 (0) | 12 (0) | 26 (0) | 0 (0) | 0 (0) | 0 (0) |
| Hepatitis | 66 (0) | 22 (0) | 45 (0) | 13 (0) | 17 (0) | 1 (0) | 0 (0) | 2 (0) |
| Hypertension | 852 (1) | 33 (0) | 673 (0) | 28 (0) | 177 (1) | 3 (1) | 0 (0) | 6 (0) |
| Hypotension | 1891 (3) | 63 (1) | 1441 (2) | 46 (1) | 970 (5) | 6 (2) | 0 (0) | 50 (2) |
| Hypothermia | 50 (0) | 3 (0) | 39 (0) | 3 (0) | 20 (0) | 0 (0) | 0 (0) | 2 (0) |
| Hypovolemia | 159 (0) | 9 (0) | 139 (0) | 7 (0) | 77 (0) | 5 (1) | 0 (0) | 0 (0) |
| Injection_site_complication | 6816 (11) | 354 (8) | 20783 (30) | 446 (13) | 1806 (10) | 28 (9) | 0 (0) | 622 (32) |

| | | | | | | | | |
|---|---|---|---|---|---|---|---|---|
| Irregular_heart_rate | 365 (0) | 62 (1) | 258 (0) | 27 (0) | 93 (0) | 0 (0) | 0 (0) | 2 (0) |
| Lymphadenopathy | 2167 (3) | 231 (5) | 2924 (4) | 134 (4) | 257 (1) | 5 (1) | 0 (0) | 22 (1) |
| Malaise | 4403 (7) | 321 (7) | 4095 (6) | 255 (8) | 1319 (7) | 23 (8) | 0 (0) | 109 (5) |
| Memory_disorder | 234 (0) | 26 (0) | 199 (0) | 28 (0) | 78 (0) | 1 (0) | 0 (0) | 1 (0) |
| Myocardial_infarction | 149 (0) | 34 (0) | 112 (0) | 19 (0) | 37 (0) | 1 (0) | 0 (0) | 1 (0) |
| Myocarditis | 49 (0) | 17 (0) | 61 (0) | 12 (0) | 11 (0) | 0 (0) | 0 (0) | 0 (0) |
| Nausea_vomiting | 7580 (12) | 456 (10) | 8323 (12) | 394 (12) | 3139 (18) | 45 (15) | 0 (0) | 159 (8) |
| Neck_back_pain | 2227 (3) | 198 (4) | 2179 (3) | 156 (4) | 862 (4) | 8 (2) | 0 (0) | 68 (3) |
| Neuropathy_NOS | 870 (1) | 69 (1) | 813 (1) | 54 (1) | 254 (1) | 2 (0) | 0 (0) | 35 (1) |
| Nonspecific_musculoskeletal_pain | 11403 (19) | 903 (20) | 14868 (21) | 747 (23) | 4260 (24) | 63 (22) | 0 (0) | 581 (30) |
| Nosebleed | 203 (0) | 19 (0) | 125 (0) | 13 (0) | 93 (0) | 2 (0) | 0 (0) | 1 (0) |
| Other_viral_infection | 974 (1) | 104 (2) | 735 (1) | 74 (2) | 172 (0) | 4 (1) | 0 (0) | 12 (0) |
| Overdose | 18 (0) | 0 (0) | 5 (0) | 0 (0) | 1 (0) | 0 (0) | 0 (0) | 0 (0) |
| Pallor | 591 (0) | 8 (0) | 504 (0) | 7 (0) | 285 (1) | 2 (0) | 0 (0) | 18 (0) |
| Pericarditis | 79 (0) | 24 (0) | 64 (0) | 12 (0) | 15 (0) | 0 (0) | 0 (0) | 0 (0) |
| Perioral_inflammation | 2944 (4) | 67 (1) | 2398 (3) | 61 (1) | 372 (2) | 5 (1) | 0 (0) | 51 (2) |
| Peripheral_neuropathy | 7066 (11) | 421 (9) | 5621 (8) | 251 (7) | 1960 (11) | 41 (14) | 0 (0) | 189 (9) |
| Peripheral_thrombosis | 2 (0) | 0 (0) | 1 (0) | 0 (0) | 2 (0) | 0 (0) | 0 (0) | 0 (0) |
| Pharyngitis | 3828 (6) | 154 (3) | 3082 (4) | 93 (2) | 607 (3) | 14 (4) | 0 (0) | 57 (3) |
| Pneumonia | 209 (0) | 41 (0) | 175 (0) | 31 (0) | 77 (0) | 1 (0) | 0 (0) | 1 (0) |
| Pregnancy_complication | 54 (0) | 23 (0) | 31 (0) | 11 (0) | 9 (0) | 1 (0) | 0 (0) | 1 (0) |
| Psychiatric_amplification | 1834 (3) | 77 (1) | 1470 (2) | 68 (2) | 444 (2) | 5 (1) | 0 (0) | 11 (0) |
| Psychiatric_depression | 150 (0) | 8 (0) | 149 (0) | 12 (0) | 54 (0) | 0 (0) | 0 (0) | 0 (0) |
| Pulmonary_embolism | 232 (0) | 67 (1) | 249 (0) | 50 (1) | 185 (1) | 7 (2) | 0 (0) | 0 (0) |
| Rash | 2377 (3) | 128 (2) | 3172 (4) | 117 (3) | 471 (2) | 13 (4) | 0 (0) | 103 (5) |
| Raynauds_phenomenon | 18 (0) | 3 (0) | 16 (0) | 0 (0) | 5 (0) | 0 (0) | 0 (0) | 1 (0) |
| Renal_failure | 85 (0) | 35 (0) | 45 (0) | 15 (0) | 19 (0) | 0 (0) | 0 (0) | 0 (0) |
| Respiratory_failure | 114 (0) | 21 (0) | 102 (0) | 12 (0) | 41 (0) | 0 (0) | 0 (0) | 0 (0) |
| Rhinitis | 1401 (2) | 123 (2) | 1054 (1) | 77 (2) | 273 (1) | 4 (1) | 0 (0) | 13 (0) |
| Seizure | 427 (0) | 23 (0) | 372 (0) | 19 (0) | 195 (1) | 4 (1) | 0 (0) | 7 (0) |
| Sepsis | 44 (0) | 7 (0) | 35 (0) | 8 (0) | 9 (0) | 0 (0) | 0 (0) | 2 (0) |
| Shock | 7 (0) | 1 (0) | 5 (0) | 1 (0) | 3 (0) | 0 (0) | 0 (0) | 0 (0) |
| Skin_infection | 55 (0) | 6 (0) | 373 (0) | 14 (0) | 17 (0) | 1 (0) | 0 (0) | 17 (0) |
| Smell_disorder | 1933 (3) | 231 (5) | 2007 (2) | 175 (5) | 872 (5) | 9 (3) | 0 (0) | 74 (3) |
| Speech_disorder | 806 (1) | 40 (0) | 607 (0) | 34 (1) | 207 (1) | 7 (2) | 0 (0) | 20 (1) |

| | | | | | | | | |
|---|---|---|---|---|---|---|---|---|
| Tachycardia | 4374 (7) | 230 (5) | 3466 (5) | 159 (4) | 879 (5) | 15 (5) | 0 (0) | 47 (2) |
| Taste_disorder | 1520 (2) | 106 (2) | 1215 (1) | 72 (2) | 308 (1) | 5 (1) | 0 (0) | 5 (0) |
| Thrombocytopenia | 84 (0) | 5 (0) | 74 (0) | 12 (0) | 37 (0) | 2 (0) | 0 (0) | 3 (0) |
| Thyroiditis | 11 (0) | 3 (0) | 5 (0) | 4 (0) | 2 (0) | 0 (0) | 0 (0) | 0 (0) |
| Tongue_pain | 953 (1) | 20 (0) | 840 (1) | 15 (0) | 118 (0) | 1 (0) | 0 (0) | 10 (0) |
| Toothache | 95 (0) | 11 (0) | 70 (0) | 14 (0) | 29 (0) | 0 (0) | 0 (0) | 2 (0) |
| Trauma | 172 (0) | 4 (0) | 148 (0) | 2 (0) | 107 (0) | 1 (0) | 0 (0) | 7 (0) |
| Urinary_frequency | 114 (0) | 7 (0) | 89 (0) | 7 (0) | 67 (0) | 1 (0) | 0 (0) | 6 (0) |
| Vaginal_infection | 2 (0) | 0 (0) | 2 (0) | 1 (0) | 1 (0) | 0 (0) | 0 (0) | 0 (0) |
| Visual_changes | 1530 (2) | 125 (2) | 1259 (1) | 88 (2) | 751 (4) | 13 (4) | 0 (0) | 27 (1) |
| Weakness | 3498 (5) | 298 (6) | 3870 (5) | 233 (7) | 1489 (8) | 19 (6) | 0 (0) | 226 (11) |
| Weight_gain | 25 (0) | 7 (0) | 23 (0) | 3 (0) | 17 (0) | 0 (0) | 0 (0) | 0 (0) |
| **65+** | | | | | | | | |
| **Adverse Events** | **Dose 1 (N = 18,895)** | **Dose 2 (N = 2,092)** | **Dose 1 (N = 34,071)** | **Dose 2 (N = 1,680)** | **Dose 1 (N = 2,669)** | **Dose 1 (N = 83)** | **Dose 2 (N = 6)** | **Dose 1 (N = 1,861)** |
| Alopecia | 25 (0) | 7 (0) | 36 (0) | 8 (0) | 1 (0) | 0 (0) | 0 (0) | 0 (0) |
| Altered_level_of_consciousness | 585 (3) | 78 (3) | 837 (2) | 51 (3) | 117 (4) | 3 (3) | 1 (16) | 44 (2) |
| Altered_sensation | 18 (0) | 0 (0) | 29 (0) | 1 (0) | 2 (0) | 0 (0) | 0 (0) | 2 (0) |
| Anaemia | 33 (0) | 11 (0) | 53 (0) | 10 (0) | 2 (0) | 0 (0) | 0 (0) | 0 (0) |
| Anaphylaxis | 65 (0) | 1 (0) | 62 (0) | 0 (0) | 6 (0) | 1 (1) | 0 (0) | 4 (0) |
| Anorexia | 446 (2) | 49 (2) | 821 (2) | 54 (3) | 72 (2) | 4 (4) | 0 (0) | 26 (1) |
| Arthropathy | 3 (0) | 0 (0) | 1 (0) | 0 (0) | 1 (0) | 0 (0) | 0 (0) | 1 (0) |
| Asthma_exacerbation | 42 (0) | 4 (0) | 57 (0) | 3 (0) | 4 (0) | 0 (0) | 0 (0) | 1 (0) |
| Autonomic_neuropathy | 1 (0) | 0 (0) | 2 (0) | 0 (0) | 0 (0) | 0 (0) | 0 (0) | 0 (0) |
| Axillary_pain | 47 (0) | 7 (0) | 116 (0) | 4 (0) | 6 (0) | 0 (0) | 0 (0) | 5 (0) |
| Bacterial_infection_NOS | 102 (0) | 16 (0) | 140 (0) | 15 (0) | 9 (0) | 0 (0) | 0 (0) | 1 (0) |
| Bleeding_NOS | 53 (0) | 2 (0) | 48 (0) | 6 (0) | 7 (0) | 1 (1) | 0 (0) | 3 (0) |
| Bowel_pattern_changes | 936 (4) | 98 (4) | 1422 (4) | 71 (4) | 125 (4) | 5 (6) | 0 (0) | 48 (2) |
| Bradycardia | 73 (0) | 8 (0) | 68 (0) | 6 (0) | 8 (0) | 1 (1) | 0 (0) | 2 (0) |
| Breast_pain | 42 (0) | 2 (0) | 62 (0) | 5 (0) | 6 (0) | 0 (0) | 0 (0) | 1 (0) |
| COPD_exacerbation | 39 (0) | 12 (0) | 41 (0) | 9 (0) | 10 (0) | 1 (1) | 0 (0) | 0 (0) |
| Cardiorespiratory_arrest | 108 (0) | 26 (1) | 115 (0) | 13 (0) | 22 (0) | 1 (1) | 0 (0) | 4 (0) |
| Central_neuritis | 4 (0) | 1 (0) | 10 (0) | 1 (0) | 0 (0) | 0 (0) | 0 (0) | 0 (0) |
| Central_neuropathy | 3003 (15) | 204 (9) | 4128 (12) | 227 (13) | 465 (17) | 14 (16) | 1 (16) | 196 (10) |
| Cerebral_hemorrhage | 49 (0) | 13 (0) | 42 (0) | 6 (0) | 17 (0) | 1 (1) | 0 (0) | 1 (0) |

| | | | | | | | | |
|---|---|---|---|---|---|---|---|---|
| Cerebral_thrombosis_infarction | 308 (1) | 59 (2) | 355 (1) | 42 (2) | 99 (3) | 2 (2) | 0 (0) | 8 (0) |
| Chest_pain | 748 (3) | 81 (3) | 858 (2) | 59 (3) | 117 (4) | 1 (1) | 1 (16) | 36 (1) |
| Choking | 25 (0) | 2 (0) | 17 (0) | 1 (0) | 0 (0) | 0 (0) | 0 (0) | 0 (0) |
| Cognitive_disorder | 245 (1) | 35 (1) | 395 (1) | 40 (2) | 48 (1) | 1 (1) | 0 (0) | 10 (0) |
| Congestive_heart_failure | 55 (0) | 13 (0) | 50 (0) | 3 (0) | 10 (0) | 0 (0) | 0 (0) | 0 (0) |
| Cough | 856 (4) | 173 (8) | 805 (2) | 104 (6) | 103 (3) | 4 (4) | 1 (16) | 24 (1) |
| Death | 870 (4) | 274 (13) | 952 (2) | 126 (7) | 151 (5) | 4 (4) | 0 (0) | 8 (0) |
| Deep_vein_thrombosis | 86 (0) | 21 (1) | 102 (0) | 23 (1) | 65 (2) | 1 (1) | 1 (16) | 0 (0) |
| Dehydration | 105 (0) | 18 (0) | 119 (0) | 5 (0) | 16 (0) | 1 (1) | 0 (0) | 6 (0) |
| Dermatitis_NOS | 1898 (10) | 116 (5) | 5890 (17) | 172 (10) | 191 (7) | 8 (9) | 0 (0) | 192 (10) |
| Dizziness | 191 (1) | 17 (0) | 248 (0) | 22 (1) | 34 (1) | 0 (0) | 0 (0) | 13 (0) |
| Drooling | 14 (0) | 0 (0) | 13 (0) | 0 (0) | 4 (0) | 0 (0) | 0 (0) | 0 (0) |
| Dry_mouth | 90 (0) | 3 (0) | 97 (0) | 1 (0) | 8 (0) | 0 (0) | 0 (0) | 5 (0) |
| Dysphagia | 214 (1) | 18 (0) | 223 (0) | 12 (0) | 31 (1) | 1 (1) | 1 (16) | 12 (0) |
| Dyspnea_NOS | 1751 (9) | 320 (15) | 1924 (5) | 224 (13) | 276 (10) | 8 (9) | 0 (0) | 61 (3) |
| Ear_pain_discomfort | 161 (0) | 14 (0) | 204 (0) | 13 (0) | 23 (0) | 1 (1) | 0 (0) | 5 (0) |
| Edema | 915 (4) | 77 (3) | 2599 (7) | 93 (5) | 162 (6) | 2 (2) | 0 (0) | 238 (12) |
| Endocrine_disorder | 68 (0) | 3 (0) | 114 (0) | 3 (0) | 10 (0) | 0 (0) | 0 (0) | 2 (0) |
| Erythema_NOS | 562 (2) | 24 (1) | 2397 (7) | 34 (2) | 61 (2) | 1 (1) | 0 (0) | 188 (10) |
| Eye_irritation | 331 (1) | 18 (0) | 473 (1) | 15 (0) | 45 (1) | 2 (2) | 1 (16) | 32 (1) |
| Facial_neuropathy | 282 (1) | 36 (1) | 313 (0) | 18 (1) | 41 (1) | 1 (1) | 0 (0) | 11 (0) |
| Failure_to_thrive | 496 (2) | 64 (3) | 691 (2) | 63 (3) | 95 (3) | 4 (4) | 0 (0) | 47 (2) |
| Fatigue | 2539 (13) | 230 (10) | 4456 (13) | 246 (14) | 390 (14) | 15 (18) | 2 (33) | 119 (6) |
| Fever | 3217 (17) | 277 (13) | 6884 (20) | 333 (19) | 556 (20) | 22 (26) | 1 (16) | 346 (18) |
| Flushing | 276 (1) | 3 (0) | 356 (1) | 4 (0) | 24 (0) | 0 (0) | 0 (0) | 8 (0) |
| Focal_musculoskeletal_pain | 87 (0) | 9 (0) | 114 (0) | 8 (0) | 15 (0) | 0 (0) | 0 (0) | 21 (1) |
| Focal_skin_abnormality | 96 (0) | 12 (0) | 161 (0) | 6 (0) | 16 (0) | 1 (1) | 0 (0) | 7 (0) |
| Gastrointestinal_bleeding | 30 (0) | 10 (0) | 19 (0) | 4 (0) | 0 (0) | 0 (0) | 0 (0) | 1 (0) |
| Groin_pain | 25 (0) | 3 (0) | 26 (0) | 3 (0) | 4 (0) | 0 (0) | 0 (0) | 0 (0) |
| Guillain_Barre_syndrome | 26 (0) | 3 (0) | 39 (0) | 8 (0) | 13 (0) | 0 (0) | 0 (0) | 17 (0) |
| Gynecologic_changes | 15 (0) | 2 (0) | 15 (0) | 2 (0) | 2 (0) | 0 (0) | 0 (0) | 0 (0) |
| Head_trauma | 3 (0) | 1 (0) | 7 (0) | 1 (0) | 1 (0) | 0 (0) | 0 (0) | 0 (0) |
| Headache | 2653 (14) | 157 (7) | 4731 (13) | 202 (12) | 478 (17) | 16 (19) | 1 (16) | 148 (7) |
| Hearing_changes | 438 (2) | 67 (3) | 471 (1) | 68 (4) | 65 (2) | 1 (1) | 0 (0) | 16 (0) |
| Hematuria | 45 (0) | 12 (0) | 43 (0) | 5 (0) | 10 (0) | 0 (0) | 0 (0) | 2 (0) |

| | | | | | | | | |
|---|---|---|---|---|---|---|---|---|
| Hepatitis | 18 (0) | 7 (0) | 28 (0) | 5 (0) | 3 (0) | 0 (0) | 0 (0) | 0 (0) |
| Hypertension | 307 (1) | 26 (1) | 366 (1) | 19 (1) | 34 (1) | 1 (1) | 0 (0) | 6 (0) |
| Hypotension | 379 (2) | 42 (2) | 462 (1) | 27 (1) | 66 (2) | 1 (1) | 0 (0) | 27 (1) |
| Hypothermia | 27 (0) | 5 (0) | 21 (0) | 0 (0) | 5 (0) | 1 (1) | 0 (0) | 1 (0) |
| Hypovolemia | 32 (0) | 1 (0) | 49 (0) | 1 (0) | 2 (0) | 0 (0) | 0 (0) | 1 (0) |
| Injection_site_complication | 1710 (9) | 75 (3) | 10021 (29) | 147 (8) | 185 (6) | 10 (12) | 0 (0) | 623 (33) |
| Irregular_heart_rate | 272 (1) | 33 (1) | 297 (0) | 35 (2) | 37 (1) | 0 (0) | 0 (0) | 7 (0) |
| Lymphadenopathy | 289 (1) | 20 (0) | 473 (1) | 35 (2) | 21 (0) | 2 (2) | 0 (0) | 11 (0) |
| Malaise | 1502 (7) | 97 (4) | 2072 (6) | 97 (5) | 213 (7) | 8 (9) | 1 (16) | 94 (5) |
| Memory_disorder | 119 (0) | 15 (0) | 143 (0) | 8 (0) | 24 (0) | 1 (1) | 0 (0) | 5 (0) |
| Myocardial_infarction | 146 (0) | 33 (1) | 152 (0) | 27 (1) | 28 (1) | 0 (0) | 0 (0) | 2 (0) |
| Myocarditis | 14 (0) | 4 (0) | 15 (0) | 5 (0) | 3 (0) | 0 (0) | 0 (0) | 0 (0) |
| Nausea_vomiting | 1966 (10) | 142 (6) | 3379 (9) | 160 (9) | 324 (12) | 12 (14) | 1 (16) | 163 (8) |
| Neck_back_pain | 668 (3) | 49 (2) | 968 (2) | 65 (3) | 110 (4) | 2 (2) | 0 (0) | 62 (3) |
| Neuropathy_NOS | 177 (0) | 8 (0) | 299 (0) | 15 (0) | 29 (1) | 0 (0) | 0 (0) | 12 (0) |
| Nonspecific_musculoskeletal_pain | 3368 (17) | 247 (11) | 6981 (20) | 301 (17) | 489 (18) | 20 (24) | 1 (16) | 520 (27) |
| Nosebleed | 76 (0) | 12 (0) | 102 (0) | 2 (0) | 18 (0) | 0 (0) | 0 (0) | 0 (0) |
| Other_viral_infection | 372 (1) | 26 (1) | 399 (1) | 31 (1) | 36 (1) | 1 (1) | 0 (0) | 12 (0) |
| Overdose | 5 (0) | 0 (0) | 3 (0) | 0 (0) | 0 (0) | 0 (0) | 0 (0) | 0 (0) |
| Pallor | 72 (0) | 2 (0) | 128 (0) | 5 (0) | 15 (0) | 0 (0) | 0 (0) | 4 (0) |
| Pericarditis | 26 (0) | 5 (0) | 20 (0) | 6 (0) | 7 (0) | 0 (0) | 0 (0) | 1 (0) |
| Perioral_inflammation | 566 (2) | 12 (0) | 689 (2) | 14 (0) | 45 (1) | 0 (0) | 0 (0) | 21 (1) |
| Peripheral_neuropathy | 1091 (5) | 64 (3) | 1433 (4) | 81 (4) | 182 (6) | 4 (4) | 0 (0) | 85 (4) |
| Peripheral_thrombosis | 2 (0) | 1 (0) | 2 (0) | 2 (0) | 7 (0) | 0 (0) | 0 (0) | 0 (0) |
| Pharyngitis | 702 (3) | 38 (1) | 866 (2) | 28 (1) | 60 (2) | 3 (3) | 0 (0) | 32 (1) |
| Pneumonia | 309 (1) | 103 (4) | 307 (0) | 72 (4) | 48 (1) | 2 (2) | 0 (0) | 3 (0) |
| Psychiatric_amplification | 477 (2) | 21 (1) | 484 (1) | 22 (1) | 53 (1) | 1 (1) | 0 (0) | 14 (0) |
| Psychiatric_depression | 34 (0) | 4 (0) | 53 (0) | 3 (0) | 12 (0) | 0 (0) | 0 (0) | 0 (0) |
| Pulmonary_embolism | 139 (0) | 51 (2) | 190 (0) | 47 (2) | 88 (3) | 4 (4) | 1 (16) | 0 (0) |
| Rash | 457 (2) | 30 (1) | 996 (2) | 29 (1) | 55 (2) | 3 (3) | 0 (0) | 55 (2) |
| Raynauds_phenomenon | 2 (0) | 0 (0) | 2 (0) | 0 (0) | 0 (0) | 0 (0) | 0 (0) | 0 (0) |
| Renal_failure | 214 (1) | 124 (5) | 142 (0) | 46 (2) | 13 (0) | 2 (2) | 1 (16) | 0 (0) |
| Respiratory_failure | 198 (1) | 73 (3) | 160 (0) | 42 (2) | 38 (1) | 1 (1) | 0 (0) | 3 (0) |
| Rhinitis | 382 (2) | 24 (1) | 477 (1) | 20 (1) | 42 (1) | 2 (2) | 0 (0) | 14 (0) |
| Seizure | 126 (0) | 17 (0) | 138 (0) | 10 (0) | 20 (0) | 1 (1) | 0 (0) | 10 (0) |

| | | | | | | | | |
|---|---|---|---|---|---|---|---|---|
| Sepsis | 84 (0) | 21 (1) | 58 (0) | 10 (0) | 12 (0) | 0 (0) | 0 (0) | 2 (0) |
| Shock | 9 (0) | 6 (0) | 10 (0) | 2 (0) | 5 (0) | 0 (0) | 0 (0) | 0 (0) |
| Skin_infection | 36 (0) | 7 (0) | 162 (0) | 6 (0) | 7 (0) | 1 (1) | 0 (0) | 26 (1) |
| Smell_disorder | 588 (3) | 57 (2) | 966 (2) | 60 (3) | 91 (3) | 5 (6) | 0 (0) | 67 (3) |
| Speech_disorder | 292 (1) | 27 (1) | 399 (1) | 27 (1) | 68 (2) | 2 (2) | 0 (0) | 14 (0) |
| Tachycardia | 748 (3) | 36 (1) | 1010 (2) | 41 (2) | 95 (3) | 3 (3) | 1 (16) | 40 (2) |
| Taste_disorder | 305 (1) | 26 (1) | 396 (1) | 20 (1) | 41 (1) | 1 (1) | 0 (0) | 2 (0) |
| Thrombocytopenia | 22 (0) | 1 (0) | 34 (0) | 4 (0) | 10 (0) | 0 (0) | 0 (0) | 0 (0) |
| Thyroiditis | 2 (0) | 0 (0) | 1 (0) | 1 (0) | 1 (0) | 0 (0) | 0 (0) | 0 (0) |
| Tongue_pain | 235 (1) | 7 (0) | 273 (0) | 4 (0) | 20 (0) | 0 (0) | 0 (0) | 5 (0) |
| Toothache | 19 (0) | 0 (0) | 50 (0) | 2 (0) | 4 (0) | 0 (0) | 0 (0) | 2 (0) |
| Trauma | 50 (0) | 7 (0) | 71 (0) | 6 (0) | 9 (0) | 0 (0) | 0 (0) | 8 (0) |
| Urinary_frequency | 37 (0) | 5 (0) | 106 (0) | 5 (0) | 12 (0) | 0 (0) | 0 (0) | 5 (0) |
| Visual_changes | 428 (2) | 32 (1) | 524 (1) | 30 (1) | 89 (3) | 3 (3) | 0 (0) | 18 (0) |
| Weakness | 1623 (8) | 199 (9) | 2531 (7) | 172 (10) | 297 (11) | 10 (12) | 1 (16) | 222 (11) |
| Weight_gain | 14 (0) | 2 (0) | 20 (0) | 0 (0) | 3 (0) | 0 (0) | 0 (0) | 0 (0) |
